# Bringing Pediatric Blood Collection Into the Home: A Parent-Administered Study of RedDrop ONE

**DOI:** 10.64898/2026.02.09.26345931

**Authors:** Timothy P. Coleman, Mason Mello, Riley Kazanjian, Maggie Kazanjian, David Olsen, Jeff Coleman, John Menna

## Abstract

Frequent blood testing is a routine but burdensome reality for many children, particularly those with chronic, rare, or medically complex conditions. Repeated clinic, hospital, and laboratory visits can disrupt family life, increase stress for children and caregivers, and limit access to timely monitoring and research participation. Despite advances in pediatric care, blood collection has remained largely tethered to in-person clinical settings. This study validates a new model: safe, effective, parent-administered pediatric blood collection performed at-home.

We evaluated the RedDrop ONE capillary blood collection device in a real-world, parent-administered home setting to determine whether non-clinical caregivers can reliably collect clinically meaningful blood samples from children without venipuncture, specialized training, or in-clinic support. Conducted under Institutional Review Board (IRB) oversight, this observational usability study enrolled 50 children aged 3–17 years across a geographically diverse U.S.-based pediatric population, including healthy and medically fragile children with chronic autoimmune and rare diseases. All study activities, including enrollment, consent, instruction, collection, and sample return, were completed remotely, reflecting real-world adoption conditions rather than controlled clinical environments.

Parents successfully collected blood samples from their children at home with high consistency, low perceived pain, and strong overall acceptance. Across collections, blood and serum volumes were sufficient and reproducible, and laboratory analysis confirmed strong analytical concordance between samples collected from two different anatomical sites, arm and leg. Parents reported high confidence using the device, short collection times, and a high likelihood of completing collections on the first attempt. Importantly, both parents and children rated the overall experience as better than expected, and parents consistently reported that the RedDrop ONE experience was superior to traditional finger-prick and needle-based venous blood draws. Parents reported minimal child discomfort and greater flexibility by avoiding in-clinic phlebotomy visits. These benefits are especially meaningful for families managing chronic or rare pediatric conditions that require repeated blood monitoring. By enabling blood collection at-home, this model reduces travel burden, scheduling constraints, and procedural anxiety while maintaining analytical reliability.

This study also demonstrated that parent-administered pediatric blood collection can support real-world clinical workflows beyond research. All samples were successfully shipped overnight at ambient temperature and processed by a CLIA-certified laboratory, supporting feasibility for remote pediatric patient monitoring and decentralized clinical trials. While lipid testing served as the representative clinical use case, the volumes and consistency achieved exceeded volume thresholds commonly required for advanced downstream applications, including proteomics, metabolomics, transcriptomics, and genomic analyses.

Taken together, these findings validate parent-administered pediatric blood collection as a practical, scalable alternative to in-clinic phlebotomy for many use cases. By shifting blood collection from the clinic to the home, this approach has the potential to reduce reliance on in-person phlebotomy, integrate seamlessly into routine pediatric care, and expand access to monitoring and research for families who face geographic, logistical, or medical barriers. For health systems, researchers, and parents alike, this study supports a future in which clinically meaningful pediatric blood collection is no longer limited by healthcare facility location but instead centered on the child and family.

## Introduction

Families caring for children with chronic or complex medical conditions often face a significant quality-of-life burden related to frequent clinic, hospital, or laboratory visits for routine blood testing. These visits can be disruptive, stressful, and time-consuming, particularly for medically fragile children and their parents. There is a critical need for validated, family-centered approaches that reduce the frequency of in-person blood draws while preserving sample integrity, analytical validity, and regulatory rigor.

The RedDrop ONE capillary blood collection device received FDA 510(k) clearance under 510(k) number K234081.^1^ It is classified as a Class II medical device, a single-use blood lancet with an integral sharps injury prevention feature. Unlike fingerstick devices that often deliver inconsistent volumes and higher pain levels, RedDrop One employs a peanut-shaped, low-profile form factor that adheres to the skin and collects blood via lancet puncture combined with a vacuum-assisted draw. The device includes integrated safety features such as automatic needle retraction, making it both user-friendly and safe for home or self-administration. In a previous clinical study, RedDrop One was tested in 100 adults aged 21 and older, who reported the device as “virtually painless,” with a median pain score of 0 on a 0–9 scale.^2^ The device successfully collected ≥150 µL of blood in 97% of cases, with an average volume of 558 µL, well above the minimum threshold for routine blood testing.^3^ Importantly, 91% of samples yielded ≥400 µL. Further validation of RedDrop ONE for accurately measuring clinical parameters comes from a direct head-to-head comparison with another FDA-cleared capillary device, the Tasso+.(Lewis et al., 2025) In a study involving 10 healthy volunteers, RedDrop ONE demonstrated excellent agreement with venous blood and Tasso+ samples for key clinical biomarkers, particularly those used in anti-doping programs like hemoglobin, reticulocyte percentage, and the OFF score.^4^ The study concluded that capillary blood collected by RedDrop ONE is analytically comparable to venous samples for common clinical parameters. This reinforces the device’s value not only for decentralized research and studies but also for high-stakes applications such as longitudinal biomarker monitoring and Athlete Biological Passports (Mahendru et al., 2020).

The RedDrop ONE device was evaluated in this study to determine its suitability for parent-administered pediatric blood sampling at-home from two sampling sites in one session (**Figure 1a-c**). The study was designed to assess whether RedDrop ONE can enable safe, low-burden collection of clinically meaningful blood volumes from children at-home without requiring specialized medical training or a clinic visit. Importantly, demonstrating suitability for pediatric home use requires more than device performance alone; it requires validation within a real-world system capable of connecting with and enrolling families, guiding parents through collection, coordinating logistics, ensuring compliant sample transport, and lab testing. By integrating RedDrop ONE into O’Ryan Health’s national logistics and informatics infrastructure, this study evaluates whether the device can be reliably deployed into households at scale, positioning RedDrop ONE not as a standalone collection tool but as part of a repeatable, family-friendly at-home blood sampling solution.

**Figure 1.**
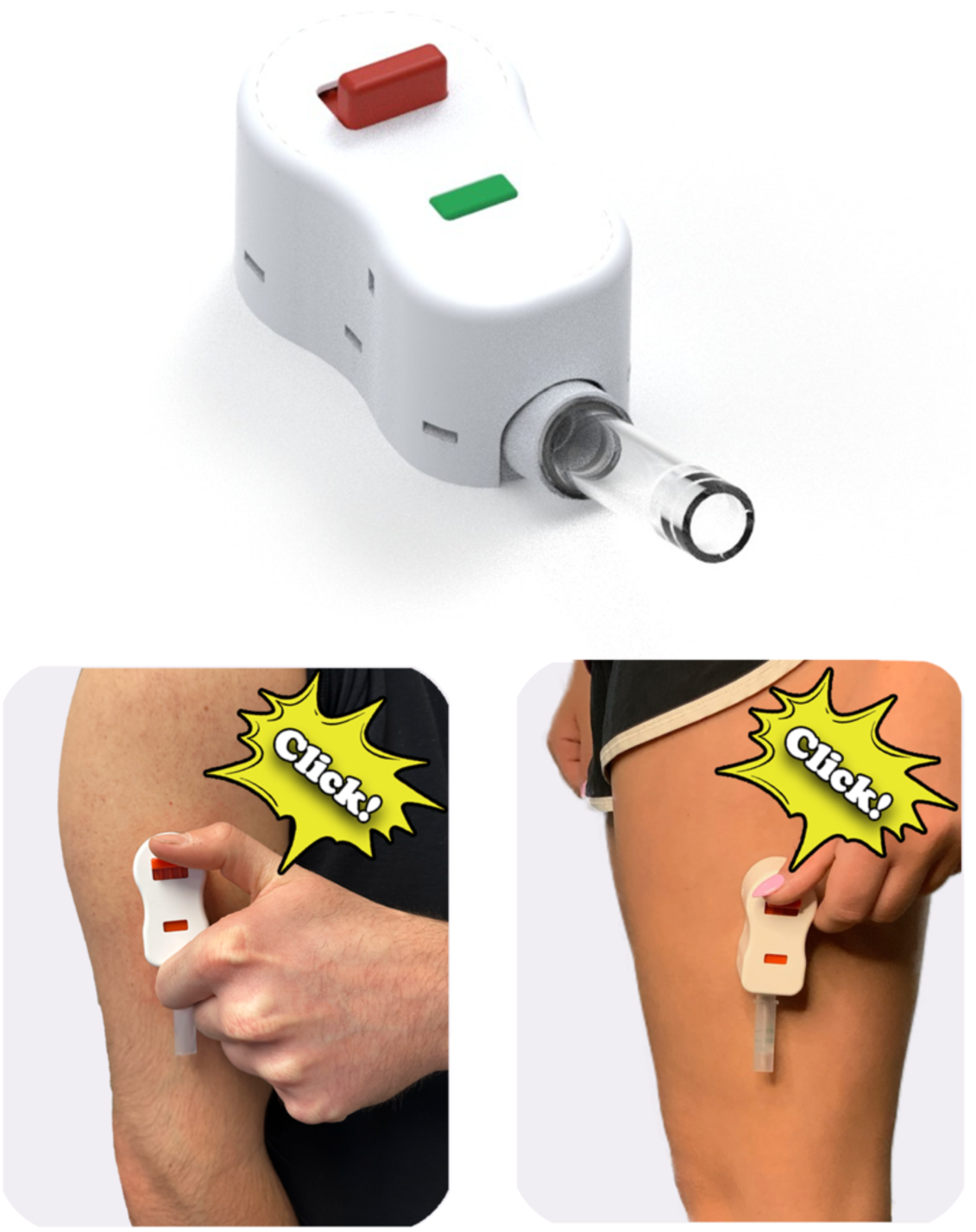
RedDrop ONE capillary blood collection device and parent-administered use. Top panel: The RedDrop ONE device with an attached microcollection tube, illustrating the integrated, vacuum-assisted capillary blood collection system evaluated in this study. Bottom panels: Representative images demonstrating parent-administered application of the RedDrop ONE device to the arm (left) and leg (right). Activation is initiated by a single button press (“click”), enabling hands-free capillary blood collection without venipuncture or specialized medical training. Images illustrate typical positioning and use under at-home conditions.

In this study, we evaluated the feasibility of caregiver-administered pediatric capillary collection using RedDrop ONE within an end-to-end remote workflow that included (i) virtual enrollment and instructions, (ii) at-home collection from 2 anatomical sites (arm and leg), (iii) overnight ambient shipping, and (iv) CLIA-certified laboratory testing. By assessing both device performance and the operational workflow required for real-world deployment, this work aims to expand access to blood-based testing for families facing geographic, logistical, or medical barriers, including those in rural communities.

## Materials and Methods

### Study Design and Setting

This study was conducted as an observational usability study evaluating the use of the RedDrop ONE device for parent-administered pediatric capillary arm and leg blood collection in real-world at-home settings. All study activities, including enrollment, consent, data collection, and sample return, were performed remotely. The study was designed to assess the feasibility, usability, and analytical performance of parent-administered blood sampling using the RedDrop ONE Device.

### Study Population and Eligibility

Children aged 2 to 17 years were eligible to participate. Enrollment was open to children with or without a known medical condition. Eligibility criteria included the parents’ willingness and ability to perform at-home blood collection using the RedDrop ONE device and access to an internet-enabled device to log in to the O’Ryan Health Parent Portal complete study surveys and instructions. Children with known contraindications to capillary blood collection, such as bleeding disorders, were excluded based on parent report.

### Recruitment

Families were recruited through multiple parent-facing channels operated by O’Ryan Health. Recruitment sources included the existing O’Ryan Health parent community, prior study participant registries, direct outreach via email and SMS, social media platforms (Instagram, Facebook, LinkedIn, X, and Discord), and public website sign-ups through a study-specific landing page. Recruitment was conducted nationwide without geographic targeting and was open to families across the United States. Enrollment was open-access and family-driven, with no targeting based on health status. Families of children with known medical conditions, as well as families of healthy children, were equally eligible to participate. Clinicians played no role in recruitment, enrollment, or study procedures.

Interested families accessed the O’Ryan Health Parent Portal, where parents completed an eligibility screening questionnaire and virtual consenting to enable enrollment for their child. Inclusion criteria required that the child be between 2.0 and 17.9 years of age at the time of enrollment, that the parent be willing and able to attempt at-home blood collection using the RedDrop ONE device, and the family have access to an internet-enabled device to complete study forms. Exclusion criteria included known bleeding disorders or other contraindications to capillary blood collection, severe anxiety around needles or blood collection, behavioral or developmental factors that would make at-home collection unsafe or unfeasible per parent judgment, or skin conditions on both the arm and leg that would interfere with safe capillary collection.

Although inclusion and exclusion criteria were defined, no children were excluded during the screening process. All families who initiated enrollment and met eligibility criteria were permitted to participate. No financial or non-financial incentives were offered for participation. Recruitment and enrollment occurred continuously between June 26, 2025, and December 18, 2025. This open-access recruitment approach was designed to reflect real-world adoption and to include families from rural, remote, and medically complex settings, consistent with the study’s goal of evaluating parent-administered pediatric blood collection in diverse home environments.

### Study Oversight and Ethical Approval

The study was approved by the Biomedical Research Alliance of New York (BRANY) Institutional Review Board, IRB File #24-10-040-1639 #253833. All parents and children >13 years old provided combined electronic informed consent before participation. Child assent was obtained when developmentally appropriate (7-12 years in age), consistent with IRB requirements. The study was conducted in accordance with applicable ethical and regulatory standards for pediatric research.

### Virtual Enrollment and Study Infrastructure

O’Ryan Health is an at-home blood sampling company purpose-built to support parent-administered blood collection for children. O’Ryan Health partners with multiple laboratories to enable general wellness programs, research studies, and CLIA-certified laboratory analysis. At the core of this capability is O’Ryan Health’s Artemis Platform, a proprietary logistics and informatics system designed to make pediatric sample collection routine, scalable, and feasible in the home. While blood collection is the primary focus of the Artemis Platform, the system is designed to support additional biospecimen types, including swabs, urine, saliva, and stool, enabling non-blood biomarker analysis and broadening the scope of pediatric research and diagnostic applications that can be conducted outside traditional clinical settings. The Artemis Platform includes an internal administrative interface that controls parent service, study configuration, and logistics. The study-level management interface enables the principal investigator to review enrollment materials and attest to complete child enrollment. The secure Parent Portal supports virtual distribution of consent, surveys, instructional content, guided sample collection, and shipping videos. All components of the Artemis Platform are intentionally designed with a consistent Superhero comic-themed user experience to maintain engagement, reduce anxiety, and center on the needs of children and families throughout participation.

Central to the Red Drop ONE study is the O’Ryan Health Parent Portal, which enables fully virtual study participation from the safety of the home. Through the Parent Portal, parents complete electronic consent, child assent (when applicable), and structured surveys to enroll their children. Once collection kits arrive at the home, the Parent Portal uses short educational videos and provides PDF instructions to guide parents through each step of the process, including pre-sample surveys, specimen collection, and return shipping.

Before the collection step, parents can view a brief instructional video on the Parent Portal demonstrating the proper use of the RedDrop ONE capillary blood collection device. Immediately before collection, parents complete a “Sample Survey” capturing information related to the child’s current general wellness, known conditions, and recent medication use at the time of sampling. After sampling, a short, second instructional video guides the packaging of the specimen and returning it via overnight UPS shipment to the designated laboratory.

### Study Device, Kit Components, and Instructions

The RedDrop ONE is a single-use capillary blood collection device designed for low-burden blood collection in non-clinical settings. The device uses a vacuum-assisted mechanism with integrated safety features, including automatic needle retraction, and is compatible with standard microcollection tubes. RedDrop ONE is FDA-cleared as a Class II medical device (510(k) K234081) and is designed to collect clinically meaningful volumes of capillary whole blood with minimal discomfort.

Each participating family received a standardized at-home collection kit, designed and distributed by O’Ryan Health Inc., containing all materials required to complete the two study blood samplings. Kit components included RedDrop ONE devices, yellow-top microcollection tubes with serum separator, alcohol swabs, gauze pads, and adhesive bandages. Two RedDrop ONE devices and corresponding supplies were allocated per anatomical site (arm and leg), with one primary and one backup RedDrop ONE device provided to ensure successful completion of study collections using a single kit.

The kit also included study-specific instructional materials designed to support parent-administered collection, including written study instructions, pictorial arm and leg sampling guides, and pictorial shipping instructions. Electronic PDF versions of these instructions, as well as videos, were available on the O’Ryan Health Parent Portal. For sample return, each kit contained a cardboard box and a pre-labeled UPS LaboratoryPak shipping container to enable overnight shipment of samples to SanoCardio. Samples were placed into the designated sample box and sealed within two labeled zip-lock plastic bags. Labels included fields for the child’s unique identifier number (UIN), time of sampling, and date of sampling to ensure accurate sample identification and processing upon receipt. All kit contents were intended to support safe, consistent, and user-friendly at-home blood collection by parents, with built-in redundancy and standardized packaging to facilitate reliable sample return and laboratory handling.

### Sample Collection Protocol

Immediately before collection, parents completed a brief sample-specific survey capturing information related to the child’s general wellness and recent health status.

Each participating child was asked to complete two capillary blood collections during a single sampling session: one from the arm and one from the leg. The order of collection sites was selected by the child, allowing for preference and comfort. A single-use RedDrop ONE device was used to collect blood from each site. During collection, parents recorded the observed fill level of the collection tube using predefined categories (first line, second line, above the second line, whole tube).

### Experience Survey

Parent and child experience with the RedDrop ONE device was assessed using a structured, study-specific Experience Survey designed to evaluate usability and associated factors during parent-administered at-home blood collection. The survey was specifically developed for this observational usability study. The Experience Survey was completed by the parent/child immediately following sample collection during the same session. Survey responses were linked to the corresponding collected sample using the child’s UIN, enabling alignment of experience metrics with blood sample lab outcomes while maintaining participant privacy.

The survey primarily assessed usability of the RedDrop ONE device and included additional questions addressing parent- and child-reported pain perception, ease of use, confidence in device operation, clarity of instructions, perceived blood volume collected, device activation and handling, and willingness to use the device again. Pain and experience measures were captured using a modified Likert Scale, a numeric rating scale ranging from 0 to 9, with anchors tailored to each question (e.g., 0 indicating no pain or much worse than expected, and 9 indicating intense pain or much better than expected)(Jebb et al., 2021). Parents reported pain and experience separately for arm and leg collection sites and provided observations regarding device performance and collection success. Parents also recorded perceived fill level of the collection tube using predefined categorical options. Optional free-text fields allowed parents to describe any issues encountered and to provide suggestions for improving the device, instructions, or overall collection process. Free-text responses were collected for qualitative feedback but were not formally coded or subjected to analysis.

Children aged 7 years and older were invited to complete an optional, age-appropriate questionnaire assessing pain, overall experience, emotional response, and willingness to use the device again. Completion of child-reported questions was voluntary, and only a subset of enrolled children provided child-reported responses. The Experience Survey also included items comparing the RedDrop ONE experience to other common blood collection experiences. These items were included as part of the overall usability assessment and were analyzed descriptively.

### Sample Handling, Shipping, and Laboratory Analysis

Following collection, samples were packaged according to the provided instructions and returned via overnight ambient-temperature shipping to a CLIA-certified clinical laboratory SanoCardio, Omaha, Nebraska. Upon receipt, laboratory personnel measured total blood volume and serum volume and performed standard lipid testing, including total cholesterol, HDL, LDL, and triglycerides. Laboratory analysis was conducted independently of all participant-facing study activities, including enrollment, consent, and at-home collection procedures.

### Outcome Measures

Primary outcomes included total blood volume, serum volume, and usability metrics related to parent-administered collection. Secondary outcomes included analytical performance of lipid measurements and concordance between arm and leg collection sites. Additional outcomes included parent- and child-reported pain, device failure rate, and willingness to use the device again.

### Statistical Analysis

Analyses were descriptive in nature. Continuous variables were summarized using medians, means, ranges, and interquartile ranges as appropriate. Categorical variables were summarized as counts and percentages. Concordance between arm and leg lipid measurements was evaluated using Pearson correlation coefficients (r) with two-sided p-values being reported. All statistical analyses and visualizations were performed using R 4.5.0.

## Results

### Geographic Reach and Enrollment Status

Virtual enrollment for the study began on June 26, 2025, and the final collected sample was received by the laboratory on December 18, 2025, representing a total enrollment and sampling period of 175 days. Across all stages of participation, families engaged with the study from a broad range of U.S. states, reflecting nationwide interest and reach (**Figure 2**). Among the 50 children who completed enrollment and sample collection, participants were distributed across 19 U.S. states, spanning the Northeast, South, Midwest, West, and non-contiguous regions. The largest concentration of enrolled participants was in Massachusetts, with additional representation across Colorado, Florida, Washington, New York, and multiple other states, including geographically remote locations such as Alaska. This distribution included urban, suburban, and diverse regional settings. In addition to the sampling cohort, seven children completed full enrollment and received at-home blood collection kits but did not complete sample collection. These participants were located across Ohio, Washington, Alaska, Massachusetts, Colorado, Florida, and Texas.

**Figure 2.**
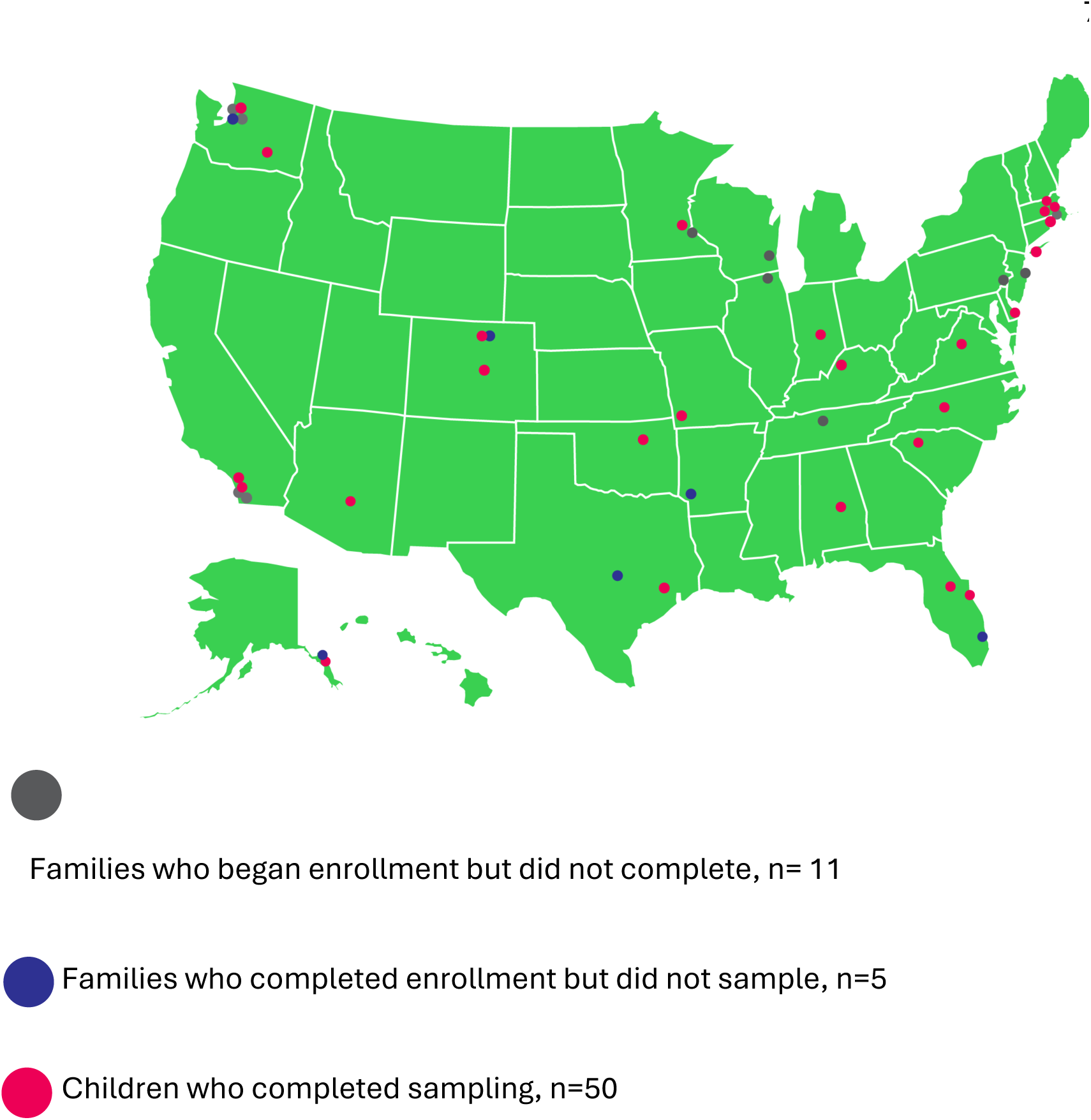
Geographic distribution and enrollment outcomes of study participants across the United States. Map illustrating the national enrollment footprint of the RedDrop ONE parent-administered pediatric blood collection study. Each marker represents a participating household, categorized by study progression status: gray dots indicate families who initiated enrollment but did not complete enrollment (*n* = 11); blue dots indicate families who completed enrollment but did not complete blood sampling (*n* = 5); and red dots indicate children who completed blood sampling (*n* = 50). The distribution demonstrates broad geographic participation across multiple U.S. states, supporting the feasibility of a decentralized, at-home pediatric blood collection model deploying RedDrop ONE at a national scale.

Further, several families initiated the enrollment process but did not complete all the required virtual steps. These initiated-but-incomplete participants were distributed across Wisconsin, Washington, Pennsylvania, Illinois, California, Tennessee, New Jersey, and Minnesota. While not included in the enrolled cohort, these families engaged with the early stages of the study workflow, including consent review and initial onboarding.

Taken together, the O’Ryan Health Artemis Platform reached families in 25 U.S. states across all stages of engagement. This geographic breadth demonstrates O’Ryan Health’s ability to distribute digital consents, coordinate nationwide logistics, and deploy the RedDrop ONE capillary blood collection device to empower parents across the United States to safely collect blood samples from their children at home for care, remote monitoring, and research participation.

### Age and Sex Distribution of Enrolled Participants

A total of 50 children participated in the RedDrop ONE study. Participant ages ranged from 3 to 17 years, reflecting inclusion across early childhood through adolescence (**Figure 3**). The median age was 13 years, with a mean age of 12 years, indicating a distribution centered in the early adolescent range. The interquartile range (IQR) spanned from 10 years (25th percentile) to 15 years (75th percentile), demonstrating that the middle 50% of participants were between late childhood and mid-adolescence. This relatively narrow IQR suggests a well-balanced age distribution without extreme skew toward younger or older participants. The cohort included 28 females and 22 males, representing a balanced sex distribution across the enrolled population. The ethnicity distribution was predominantly White or Caucasian (n = 40), with additional representation from Asian or Pacific Islander (n = 3), Asian Indian (n = 2), Black or African American (n = 2), Hispanic or Latino (n = 2), American Indian or Alaska Native (n = 1), and multiracial (Caucasian/Asian; n = 1) participants. Overall, the study population reflects a broad pediatric age, sex, and ethnic spectrum, supporting the evaluation of parent-administered blood collection across multiple developmental stages.

**Figure 3.**
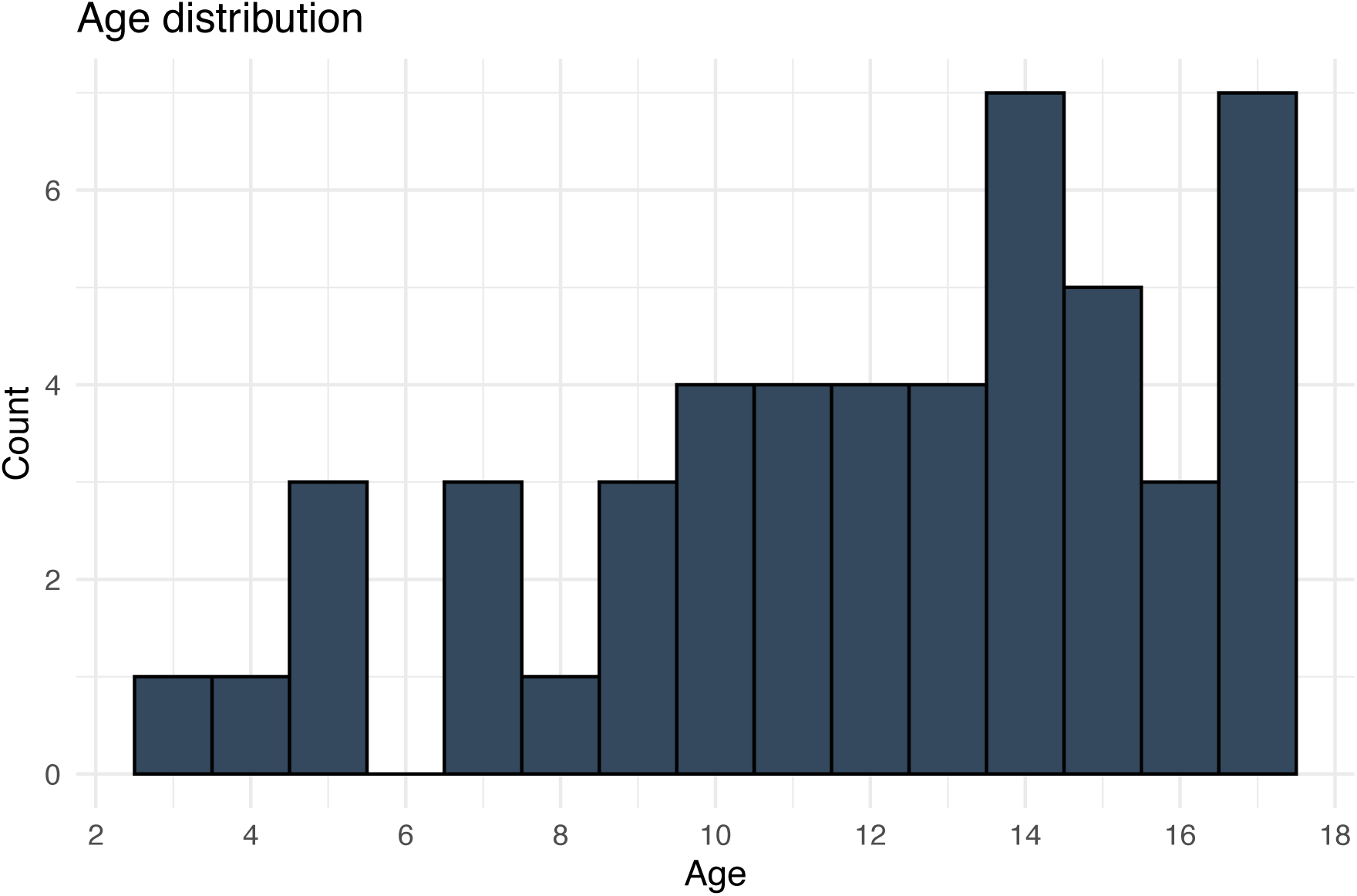
Age distribution of pediatric participants. Histogram showing the age distribution of children enrolled in the RedDrop ONE study. Participants ranged from early childhood through late adolescence, with representation across the full eligible age spectrum. The distribution supports the evaluation of RedDrop ONE usability and performance across a broad pediatric population.

### Clinical Conditions Represented in the Enrolled Cohort

Among the 50 participating children, 14 participants reported one or more underlying medical conditions, while the remaining 36 children were reported as healthy, with no known diagnosed disorders at the time of enrollment. Several of the affected participants had multiple co-existing diagnoses. The most reported condition was juvenile dermatomyositis (JDM), present in 7 children, including cases with associated autoimmune and connective tissue disorders. Reported comorbidities among children with JDM included postural orthostatic tachycardia syndrome (POTS), Hashimoto’s thyroiditis, Raynaud’s phenomenon, Sjögren syndrome, and scleroderma.

Additional conditions included Hashimoto’s thyroiditis in 4 children (including both isolated and comorbid cases) and PANS/PANDAS in 2 children. Sjögren syndrome and scleroderma were each reported in 2 children, occurring in the context of multisystem autoimmune disease. Single cases were reported for celiac disease, methylmalonic acidemia, eczema, and keratosis pilaris. Overall, the cohort comprised mostly of healthy children alongside a clinically diverse subset with autoimmune, inflammatory, metabolic, and dermatologic conditions, supporting evaluation of parent-administered blood collection across both healthy and medically complex pediatric populations.

### Sample Collection

Parent- and child-reported experience measures indicated high overall acceptability of the RedDrop ONE device, with nearly all participating families expressing willingness to use the device again for at-home blood collection. Parent confidence in device use was high, and collections were generally completed on the first attempt with minimal observable discomfort for children, supporting the usability of the device in real-world home settings.

Total blood volume and serum volume collected from arm and leg capillary sampling sites were comparable across participants, with overlapping distributions and similar central tendencies observed between collection sites, (**Figure 4**). Importantly, 95% of samples were greater than 400 μL blood. For arm-based collections using the RedDrop ONE device, total blood volume ranged from 300 to 950 µL, with a mean volume of 669 µL and a median of 650 µL. The interquartile range (25th–75th percentile) for arm collections was 600–780 µL. Leg-based collections yielded total blood volumes ranging from 360 to 1,000 µL, with a mean volume of 659 µL and a median of 610 µL. The interquartile range for leg collections was similarly narrow at 598–750 µL. The narrow and highly overlapping interquartile ranges indicate low variability and consistent volume yield across participants and collection sites, demonstrating that most collections produced reliable and reproducible blood volumes independent of puncture site. Despite slightly higher median and mean volumes observed for arm collections, no significant difference in total blood volume was observed between puncture sites.

**Figure 4.**
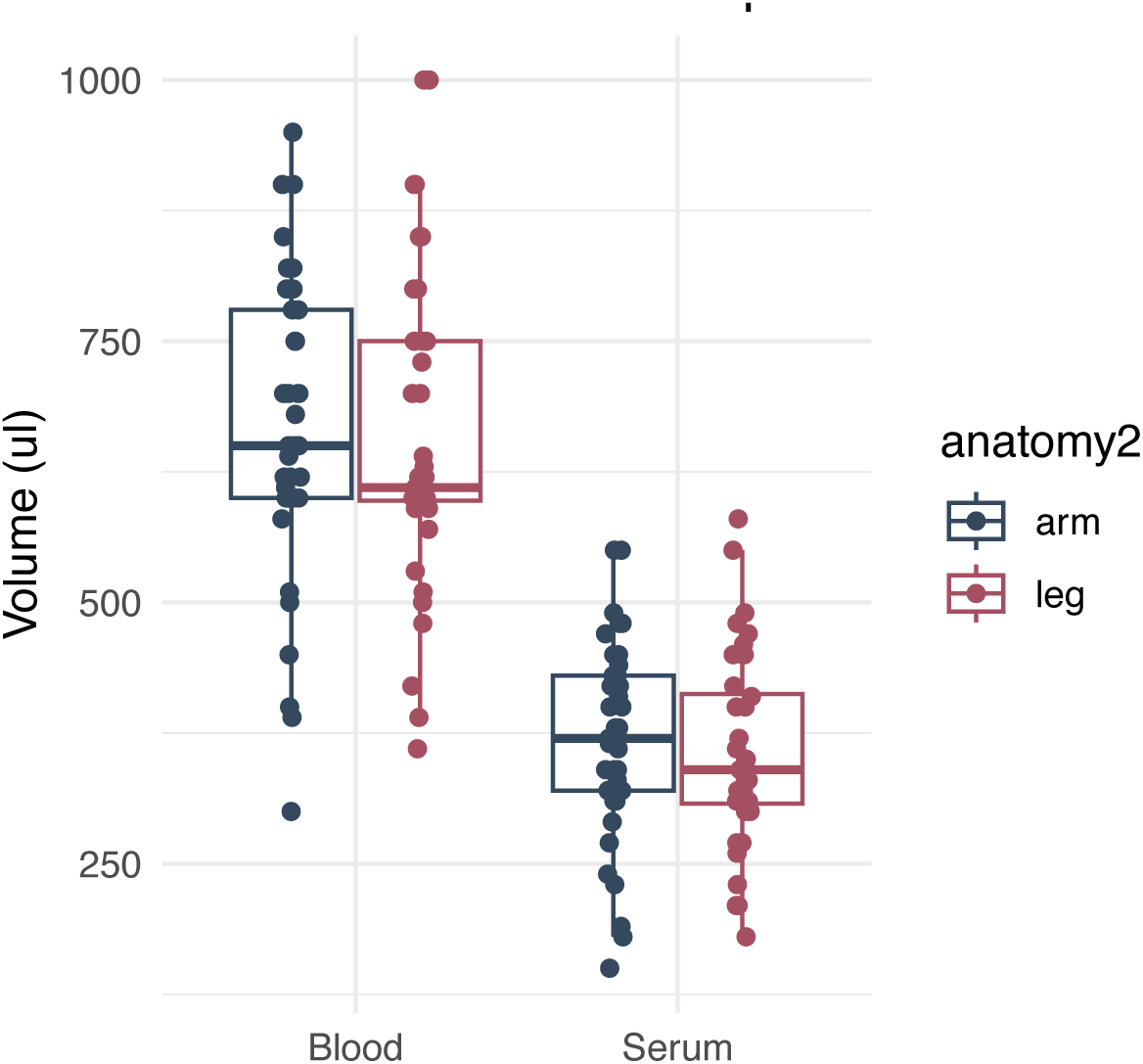
Whole blood and serum volumes collected by anatomical sampling site. Box-and-jitter plots showing whole blood and serum volumes obtained using the RedDrop ONE device from arm and leg capillary sampling sites. Individual points represent samples collected under parent-administered, at-home conditions, with box plots indicating median and interquartile ranges. Comparable distributions and overlapping central tendencies were observed between arm and leg collection sites for both whole blood and serum volumes, demonstrating that multiple anatomical sites can reliably yield clinically meaningful sample volumes in a home-based pediatric setting.

Across all collections, the median total blood volume was 615 µL (25th–75th percentile: 600–750 µL), with a mean volume of 639 µL well in accordance with prior RedDrop ONE studies performed in adults(Collier et al., 2025; Lewis et al., 2025). Median serum volume was 340 µL (25th–75th percentile: approximately 300–380 µL), with a mean of 341 µL. The relatively narrow and overlapping interquartile ranges for both total blood and serum volumes indicate low variability and consistent performance across participants and collection sites, supporting the reliability of the parent-administered collection process. Together, these findings demonstrate that both arm and leg capillary collection sites reliably generate sufficient and reproducible total blood and serum volumes for clinical lipid testing, supporting flexibility in site selection for parent-administered pediatric blood collection.

### Experience Survey

#### Parent-reported pain and anatomical site scores

Parent-reported pain scores for both arm- and leg-based capillary blood collection using the RedDrop ONE device were low overall (**Figure 5a-b**). Using a modified Likert scale (Jebb et al., 2021), a numeric rating scale from 0 (no pain) to 9 (intense pain), arm-based pain scores ranged from 0 to 7, with an average score of 2.32 and a median of 2.00, while leg-based pain scores ranged from 0 to 9, with an average score of 2.34 and a median of 1.5. These values indicate generally mild perceived discomfort across both collection sites. For both arm and leg collections, pain score distributions were skewed toward the lower end of the scale, with a substantial proportion of responses clustered between 0 and 2. Higher pain scores were infrequent and limited to a small subset of responses, with no clustering at the upper end of the scale. Overall, these findings demonstrate that parent-administered capillary blood collection using the RedDrop ONE device was associated with low perceived pain in children across both anatomical sites, with leg-based collection perceived as slightly less painful based on the lower median score, supporting the acceptability of both sites for pediatric at-home blood collection.

**Figure 5.**
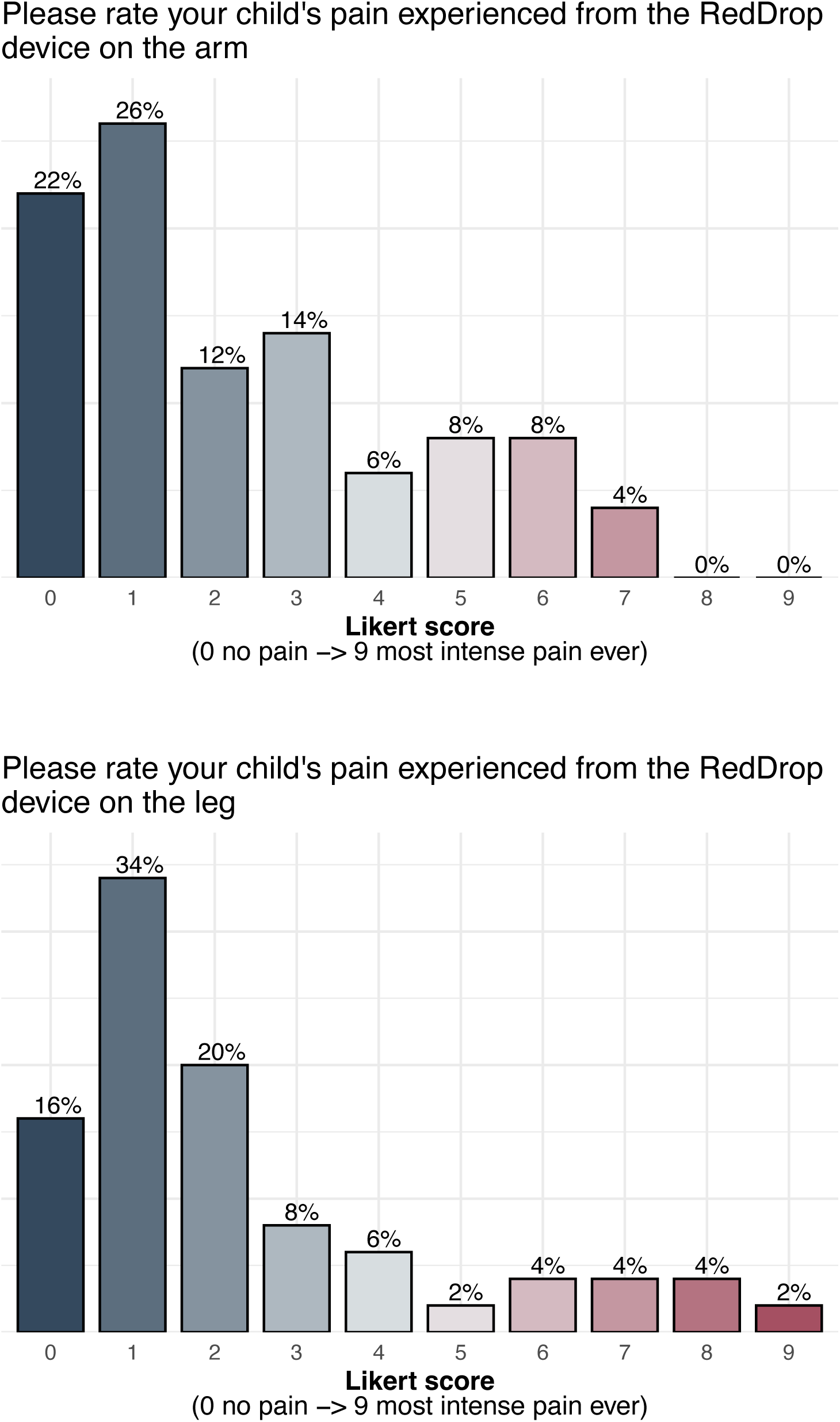
Parent-reported pain scores associated with RedDrop ONE capillary blood collection. **(a)** Distribution of parent-reported pain scores for arm-based sampling. **(b)** Distribution of parent-reported pain scores for leg-based sampling. Pain was assessed where 0 indicates no pain and 9 indicates the most intense pain ever experienced. Bars represent the percentage of responses at each pain score. For both anatomical sites, the majority of parents reported low pain scores (0–3), with progressively fewer reports at higher pain levels, indicating generally low perceived discomfort associated with parent-administered RedDrop ONE use in the home setting. Which site was easier to use the RedDrop device on?

When parents were asked which sampling site, arm or leg, was easier to use for RedDrop ONE collection, responses indicated no strong overall preference between the arm and leg (**Figure 6**). Forty-six percent (44%) of parents reported no difference in ease of use between sites, representing the largest single response category. Thirty percent of parents indicated that the leg was easier to use, while twenty-six percent of parents reported that the arm was easier to use. The predominance of “no difference” responses suggests that parents were able to use the RedDrop ONE device effectively on both sites with comparable ease. The relatively balanced distribution between arm and leg preferences further supports the usability of both collection sites and indicates that site selection can be guided by child comfort or situational factors rather than technical difficulty. Overall, these findings reinforce the flexibility of the RedDrop ONE device for parent-administered pediatric blood collection, with no clear usability disadvantage associated with either the arm or leg.

**Figure 6.**
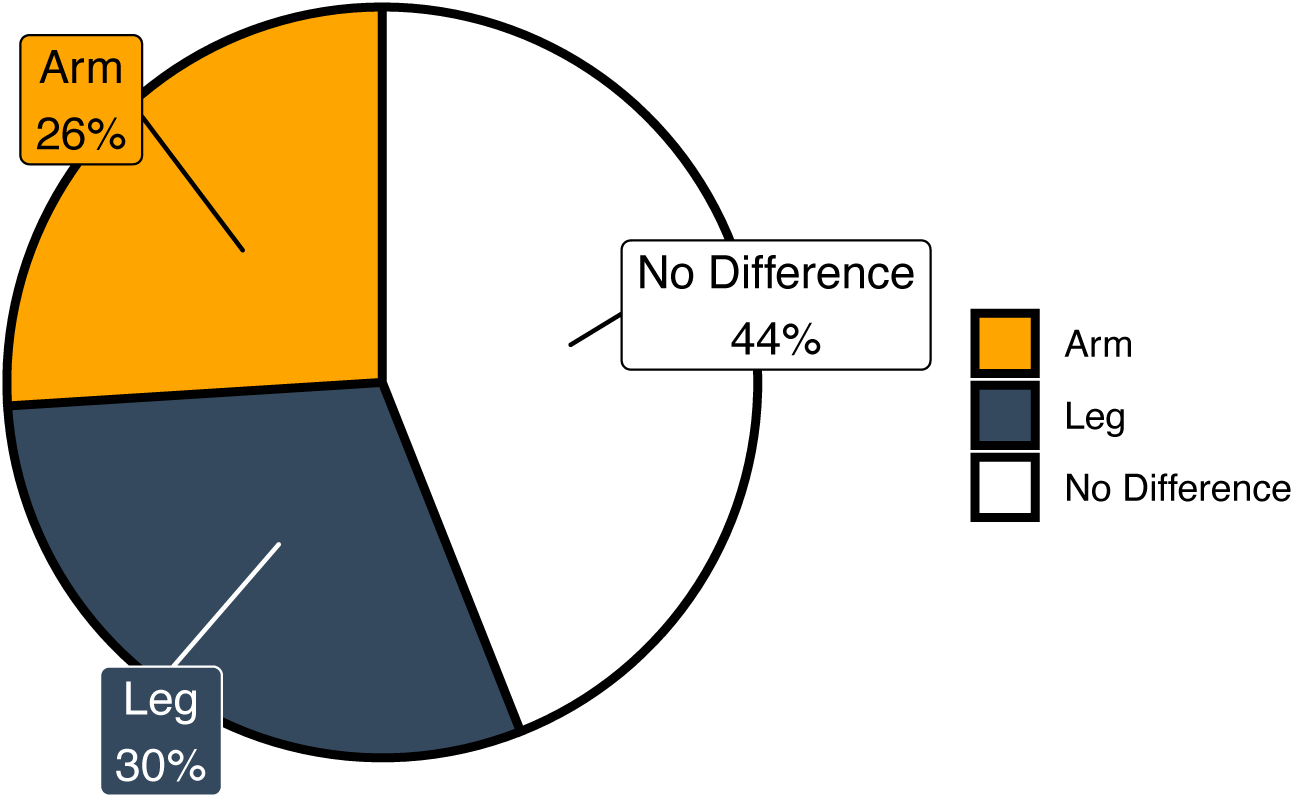
Parent-reported ease of use by anatomical sampling site. Pie chart summarizing parent responses to the question, “Which site was easier to use the RedDrop ONE device on?” Responses indicate that 44% of parents reported no difference in ease of use between arm and leg sampling sites, 30% reported the leg as easier to use, and 26% reported the arm as easier to use. These findings suggest comparable usability of RedDrop ONE across anatomical sites, with no strong preference favoring either arm or leg for parent-administered, at-home blood collection.

#### Overall Experience

Parent-reported ratings of children’s overall experience with the RedDrop ONE device were highly positive (**Figure 7**). On a numeric rating scale from 0 = worse than expected to 9 = better than expected, overall experience scores ranged from 5 to 9, with an average score of 8.26 and a median score of 9.00, indicating a strong, favorable assessment across participants. The distribution of responses was skewed toward the upper end of the scale, with most ratings clustered at 8 and 9. Lower scores were infrequent and limited to a small subset of responses, with no evidence of widespread dissatisfaction. This high concentration of top-end ratings suggests that most children had a positive experience with the device from start to finish. Overall, these findings demonstrate high parent-reported satisfaction with the RedDrop ONE experience and support its acceptability for pediatric at-home blood collection.

**Figure 7.**
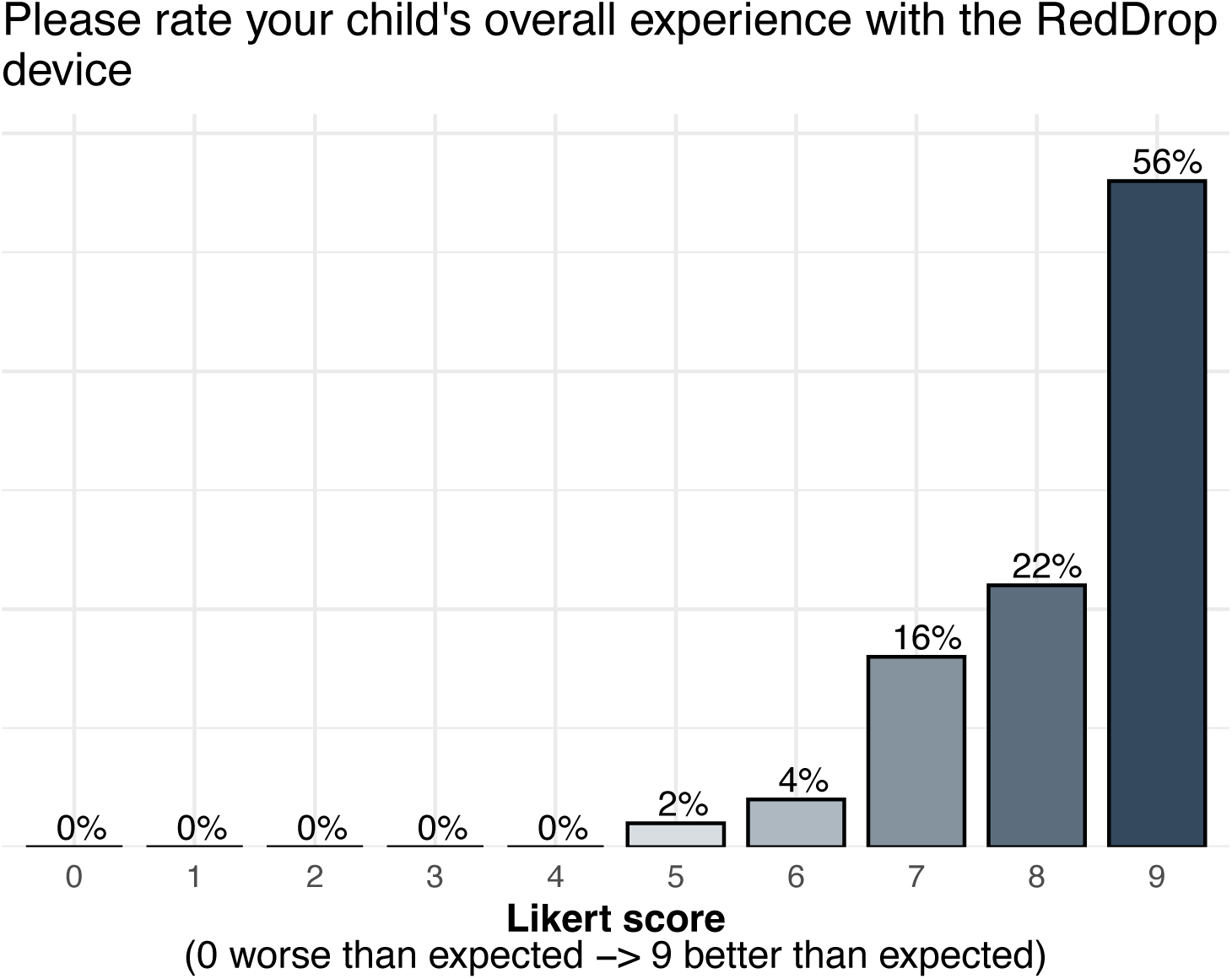
Parent-reported overall experience with RedDrop ONE. Bar chart showing parent ratings of their child’s overall experience with the RedDrop ONE device where 0 indicates worse than expected and 9 indicates better than expected. The majority of responses clustered at the high end of the scale, with most parents rating the experience as 7–9. These results indicate a strongly positive overall experience with parent-administered RedDrop ONE blood collection in the home setting.

#### Usability Outcomes Completion and Time

Most parents were able to complete blood collection on the first attempt using the RedDrop ONE device. Of the 50 respondents, 47 parents reported completing the collection on the first attempt, while 3 parents indicated that a second attempt was required, demonstrating a high first-attempt success rate. For the three failed sampling attempts, two were resolved by hydrating the child and resampling 30 minutes later, and the third failure was due to the device being improperly placed on the leg without ensuring the tube was pointed down as described in the instructions. The duration of the blood collection process (i.e., heat-pack application, blood draw, and sample packaging) was generally short. Based on parent-reported estimates, the mean collection time was approximately 10 minutes, with a median time of 8.5 minutes, indicating that most collections were completed efficiently. These findings support the feasibility of parent-administered blood collection in home settings and demonstrate that the RedDrop ONE device can be used successfully with minimal time burden on families.

#### Observed Child Comfort and Tolerance

Visible discomfort in children during or after collection was *<u>reported infrequently</u>*. Thirteen parents observed visible discomfort, while 37 parents reported no visible discomfort, indicating that most children tolerated the procedure well. Visible discomfort was not associated with failure to complete the collection on the first attempt. Overall, these findings demonstrate that parent-administered blood collection using the RedDrop ONE device can be completed successfully on the first attempt in most cases, within a short duration, and with minimal observable discomfort for children, further supporting the usability and acceptability of the device in pediatric home settings.

#### Parent Confidence and Collection Experience

Parents reported high levels of confidence when using the RedDrop ONE device to collect blood from their children. On a numeric rating scale from 0 (not at all confident) to 9 (extremely confident), confidence scores ranged from 5 to 9, with an average score of 8.26 and a median score of 9.00, indicating strong overall confidence in device use (**Figure 8**). Most parents reported no issues or difficulties during the collection process. Specifically, 40 parents indicated that no issues were encountered, while 10 parents reported experiencing some form of difficulty.

**Figure 8.**
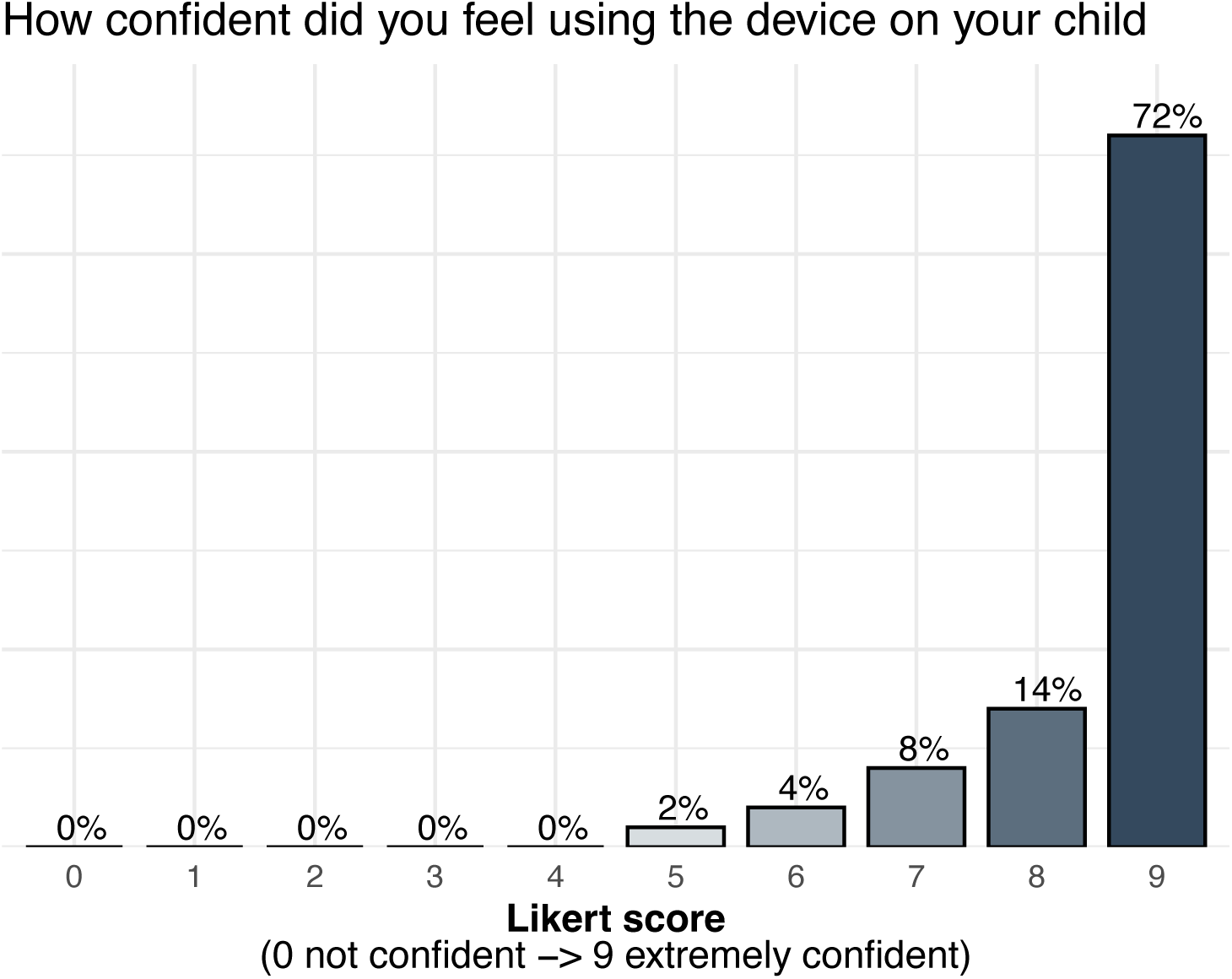
Parent-reported confidence in using the RedDrop ONE device. Bar chart showing parent-reported confidence levels associated with use of the RedDrop ONE device on their child. Confidence was assessed where 0 indicates not confident and 9 indicates extremely confident. Bars represent the percentage of responses at each confidence level. Responses were strongly skewed toward the upper end of the scale, with most parents reporting high confidence (scores 8–9) using the RedDrop ONE for at-home capillary blood collection.

Despite these reported issues, confidence scores remained high overall, suggesting that minor challenges did not substantially affect parents’ ability to complete the collection. Together, these findings demonstrate that parents felt highly confident using the RedDrop ONE device and were generally able to complete at-home blood collection with minimal difficulty, supporting the usability and reliability of the device in real-world pediatric at-home settings.

#### Parent-Reported Experience Relative to Traditional Collection Methods

Parents reported favorable comparisons between the RedDrop ONE device and traditional blood collection methods. When asked to compare the RedDrop ONE experience to a traditional finger prick using a numeric rating scale from 0 = worse than expected to 9 = better than expected, had an average of 7.92 and a median of 9.00, indicating that most parents perceived the RedDrop ONE device as better than or comparable to finger prick collection (**Figure 9**). Comparisons to a traditional needle blood draw were similarly positive. Ratings for this comparison yielded a higher average score of 8.6 and a median of 9.00, reflecting a strong preference for the RedDrop ONE experience relative to a traditional needle-based blood draw (**Figure 10**). Scores were clustered toward the upper end of the scale for both comparison questions, with relatively few low ratings, suggesting broad consistency in parent perceptions. These favorable experiences translated into a strong stated willingness to participate in future studies using the RedDrop ONE device. When asked whether they would be willing to participate in a future study using the RedDrop ONE, 82% of respondents answered “Yes,” and 14% responded “Maybe,” (**Figure 11**). Together, these findings indicate that parents generally viewed the RedDrop ONE device as an improved, more acceptable alternative to traditional pediatric blood collection methods and support its suitability for repeated, at-home use from a parent’s perspective.

**Figure 9.**
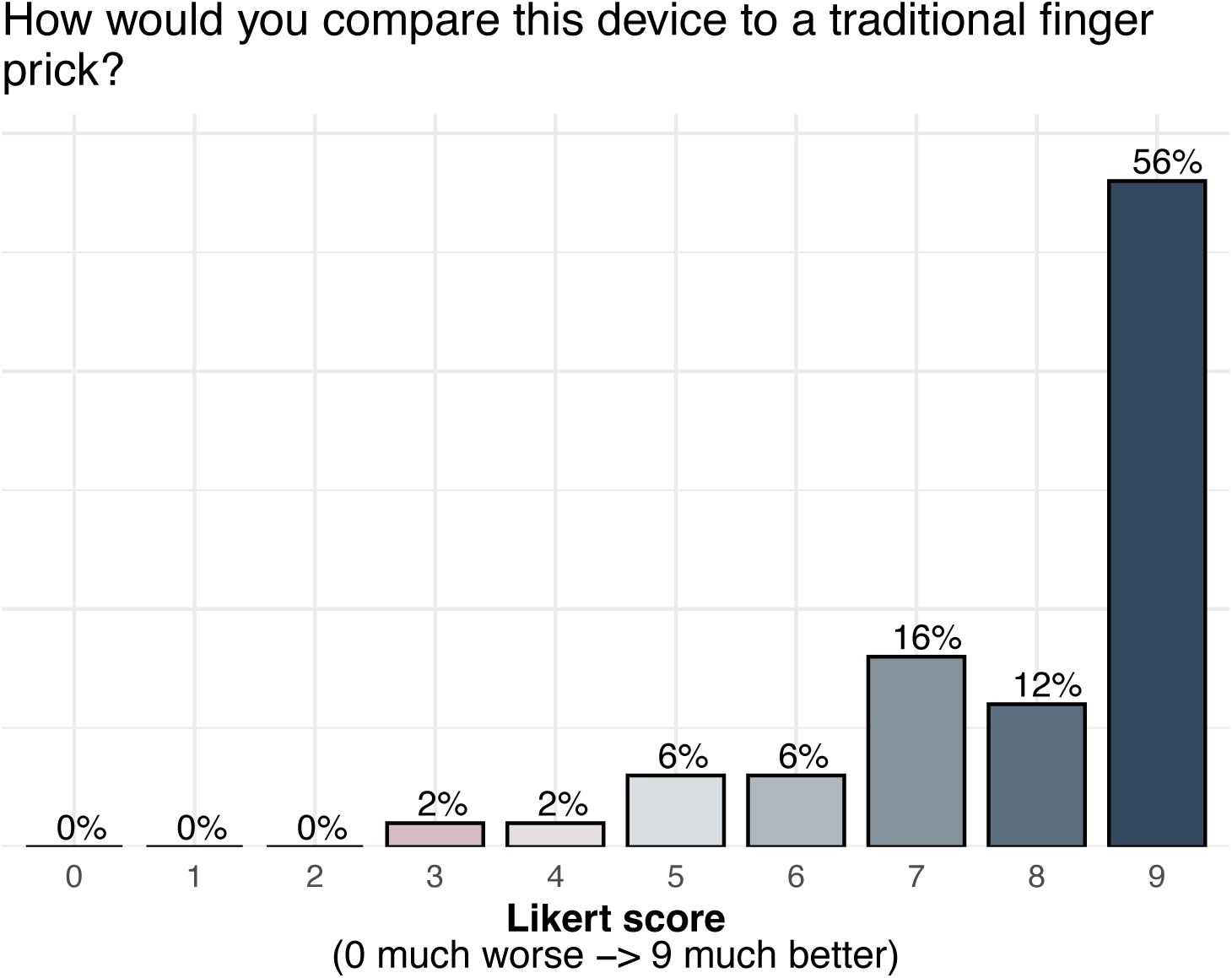
Parent-reported comparison of RedDrop ONE to traditional finger-prick blood collection. Bar chart showing parent-reported comparisons of the RedDrop ONE device relative to traditional finger-prick blood collection. Responses where 0 indicates much worse than a finger prick and 9 indicates much better than a finger prick. Bars represent the percentage of responses at each score. Ratings were predominantly distributed toward the upper end of the scale, indicating that most parents perceived RedDrop ONE as better than traditional finger-prick.

**Figure 10.**
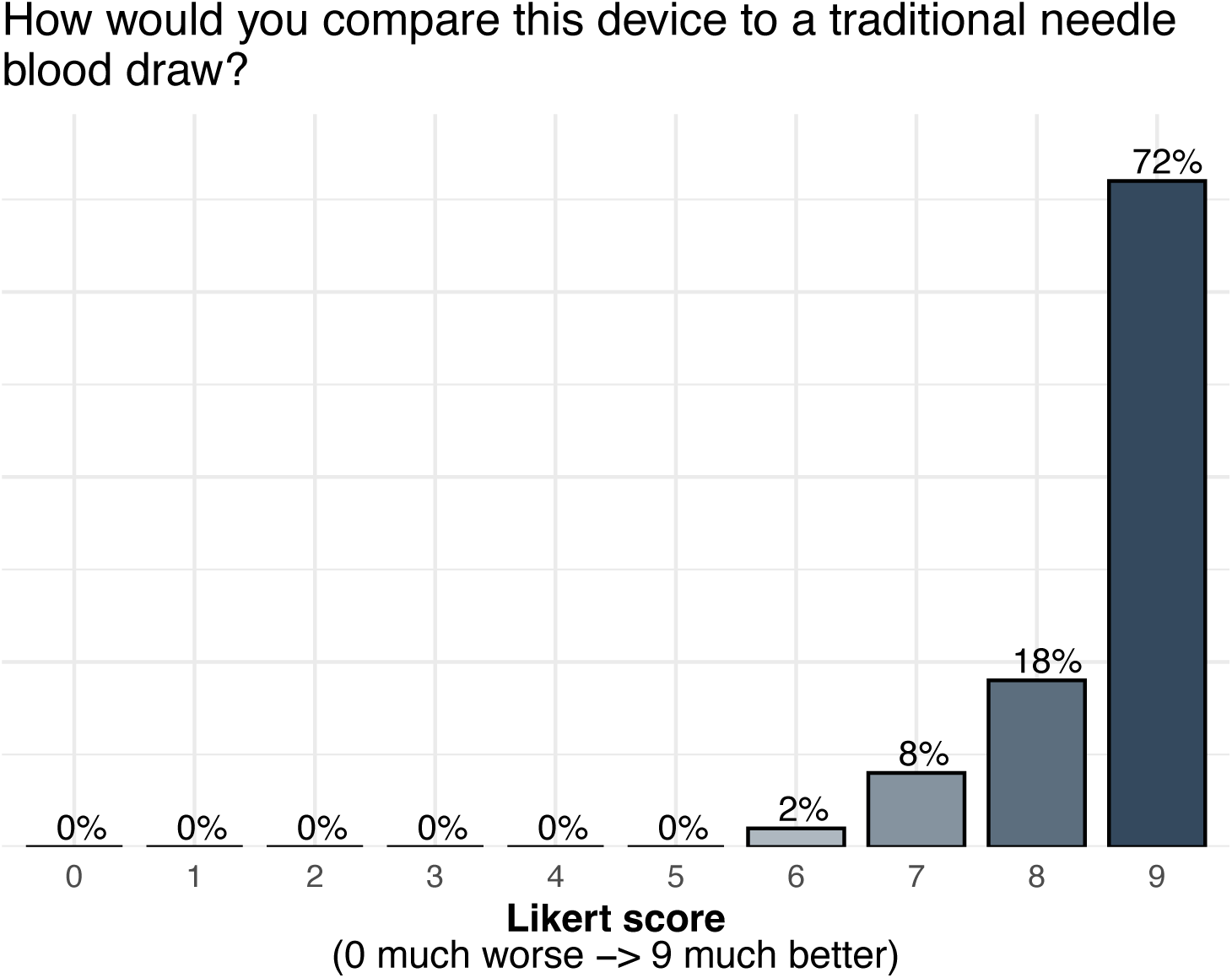
Parent-reported comparison of RedDrop ONE to traditional venous blood draw. Bar chart showing parent-reported comparisons of the RedDrop ONE device relative to traditional needle-based venous blood draw. The scale, 0 indicates much worse than a traditional blood draw and 9 indicates much better than a traditional blood draw. Bars represent the percentage of responses at each score. Responses were strongly skewed toward higher scores, indicating that most parents perceived RedDrop ONE as more favorable than conventional venipuncture.

**Figure 11.**
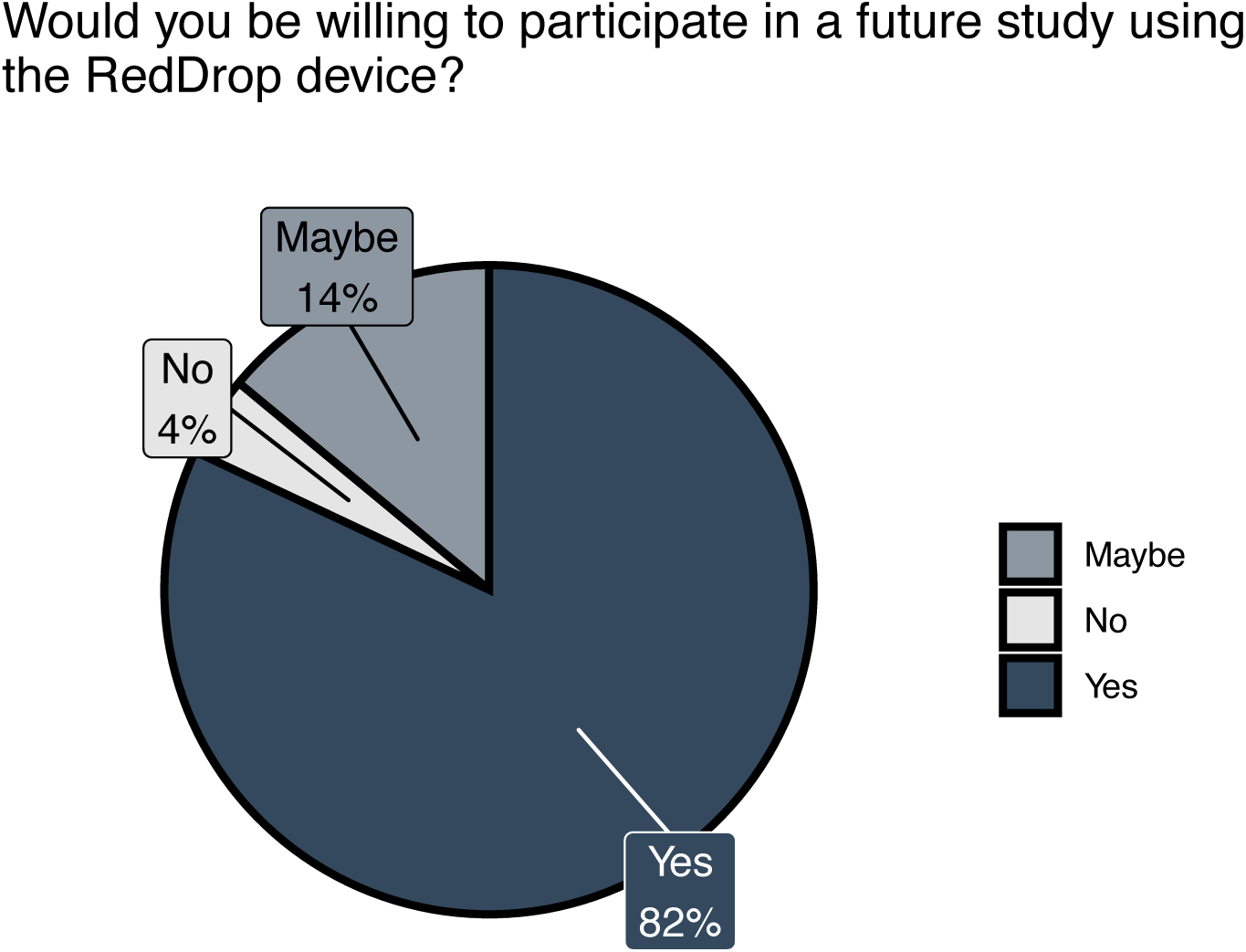
Willingness to participate in future studies using the RedDrop ONE device. Pie chart summarizing parent responses to the question, “Would you be willing to participate in a future study using the RedDrop ONE device?” Responses are shown as proportions of total respondents, with categories indicating “Yes,” “Maybe,” and “No.” The majority of parents indicated willingness to participate in a future study, with smaller proportions reporting uncertainty or unwillingness. This distribution reflects overall participant receptivity to repeat use of the Red Drop ONE in the context of the study.

#### Parent experiences with the kits

Across all kit components, parent-reported experiences were consistently positive, indicating high usability and acceptability of the RedDrop ONE system. Heat pack use was well tolerated, with the majority of parents reporting minimal or no discomfort for their child and only a small minority reporting higher discomfort (**Figure 12**). RedDrop ONE activation was rated as highly intuitive, with most participants selecting the highest possible scores and very few indicating any difficulty, (**Figure 13**). Removal of the device from the skin was similarly well received, with responses concentrated at the favorable end of the scale and no reports of poor tolerance (**Figure 14**). In parallel, comprehension of the RedDrop instructions was strong, with most participants indicating that the instructions were easy to understand and only a small number reporting moderate difficulty (**Figure 15**). Collectively, these results demonstrate that the RedDrop ONE kit components and instructions can be used comfortably and effectively by parents in a home setting, supporting the overall feasibility of parent-administered pediatric blood collection.

**Figure 12.**
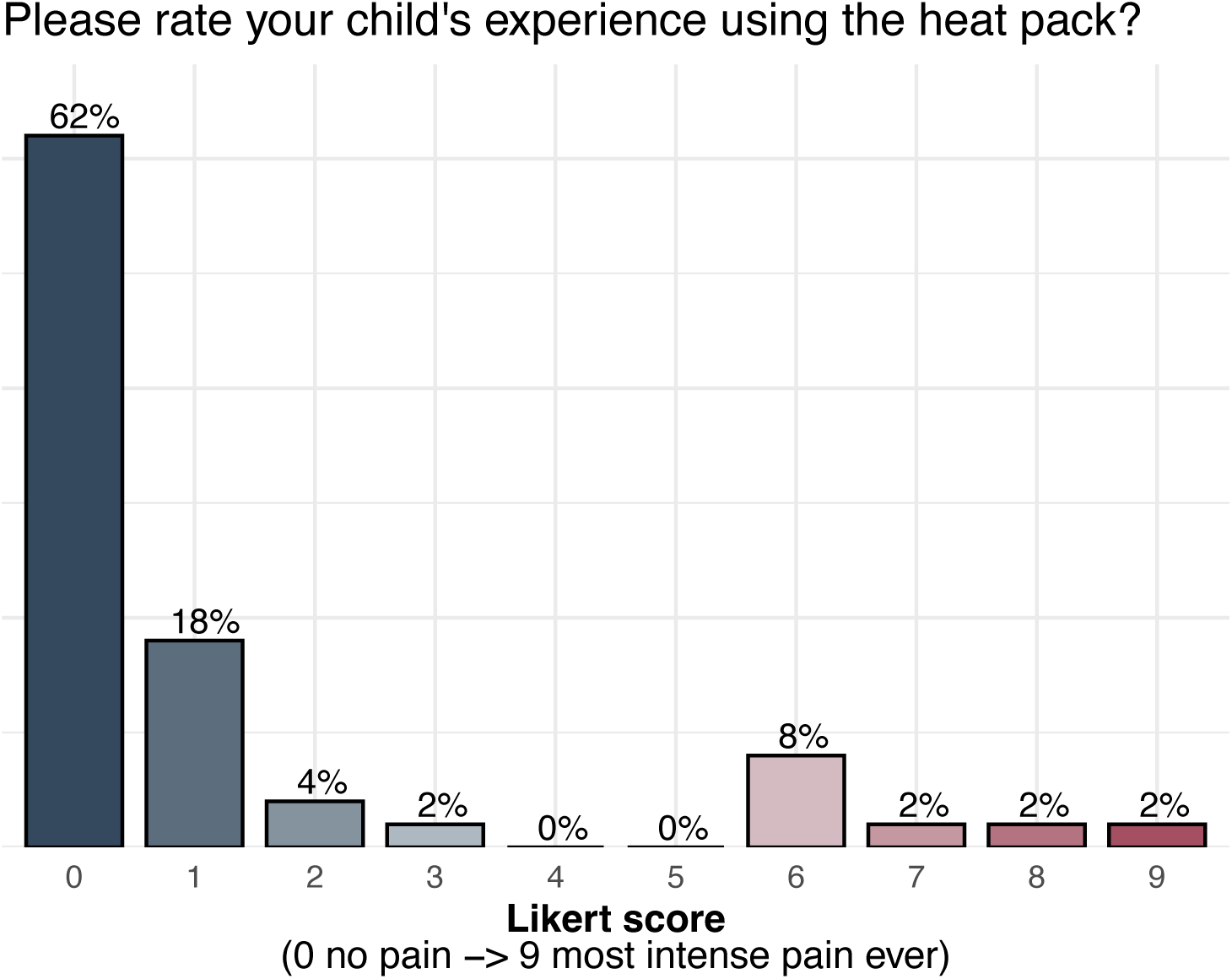
Parent-reported child experience associated with use of a warming heat pack. Bar chart showing parent-reported ratings of their child’s experience using the heat pack applied prior to RedDrop ONE capillary blood collection. Experience was assessed using the scale were 0 indicates no pain and 9 indicates the most intense pain ever experienced. Bars represent the percentage of responses at each score. Most responses clustered at the lower end of the scale, indicating minimal discomfort associated with heat pack use in the at-home setting.

**Figure 13.**
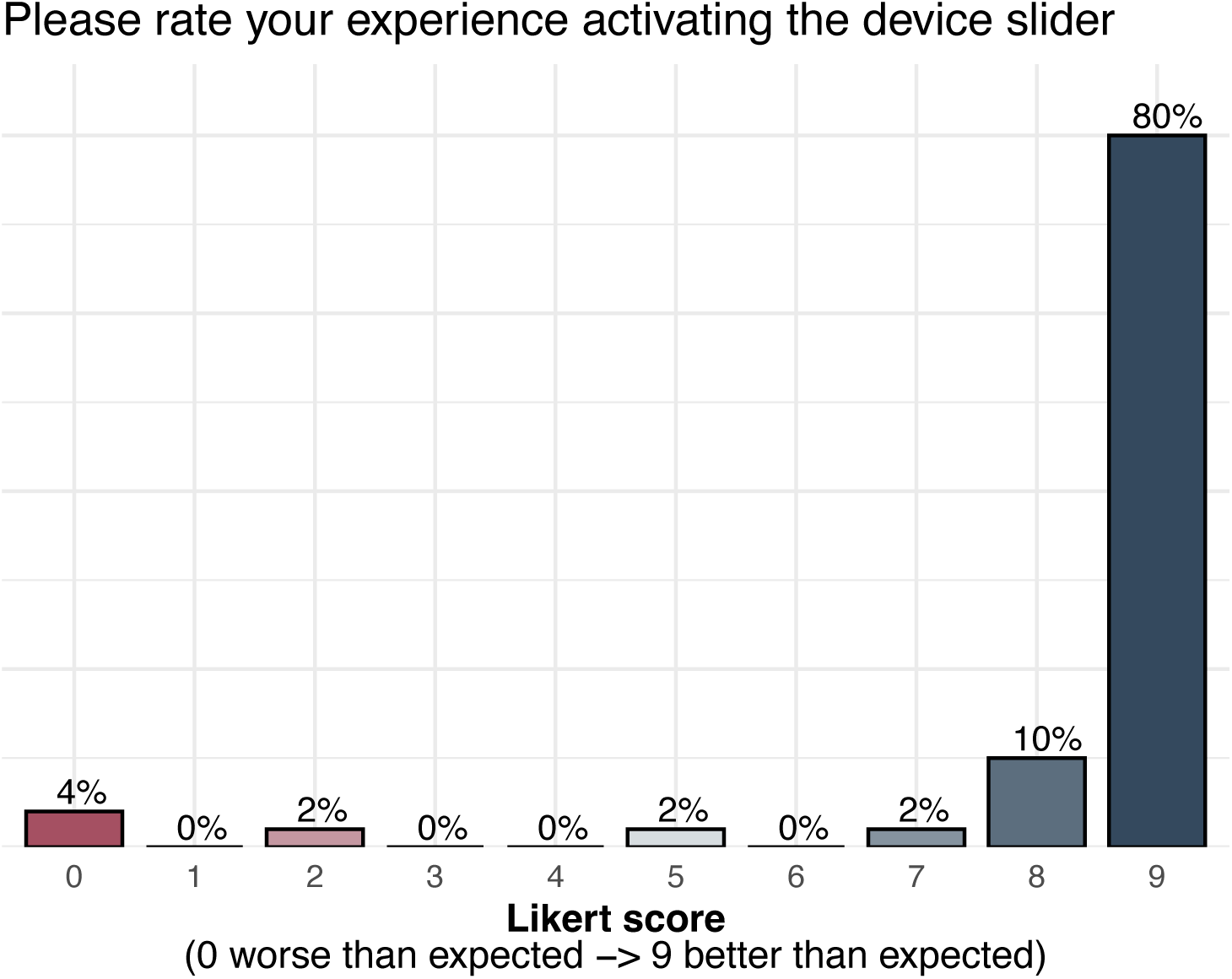
Parent-reported experience activating the RedDrop ONE device slider. Bar chart showing parent-reported ratings of their experience activating the RedDrop ONE device slider. Experience was assessed using the scale, where 0 indicates worse than expected and 9 indicates better than expected. Bars represent the percentage of responses at each score. Responses were strongly skewed toward higher ratings, indicating that most parents found RedDrop ONE device activation intuitive and favorable during parent-administered, at-home capillary blood collection.

**Figure 14.**
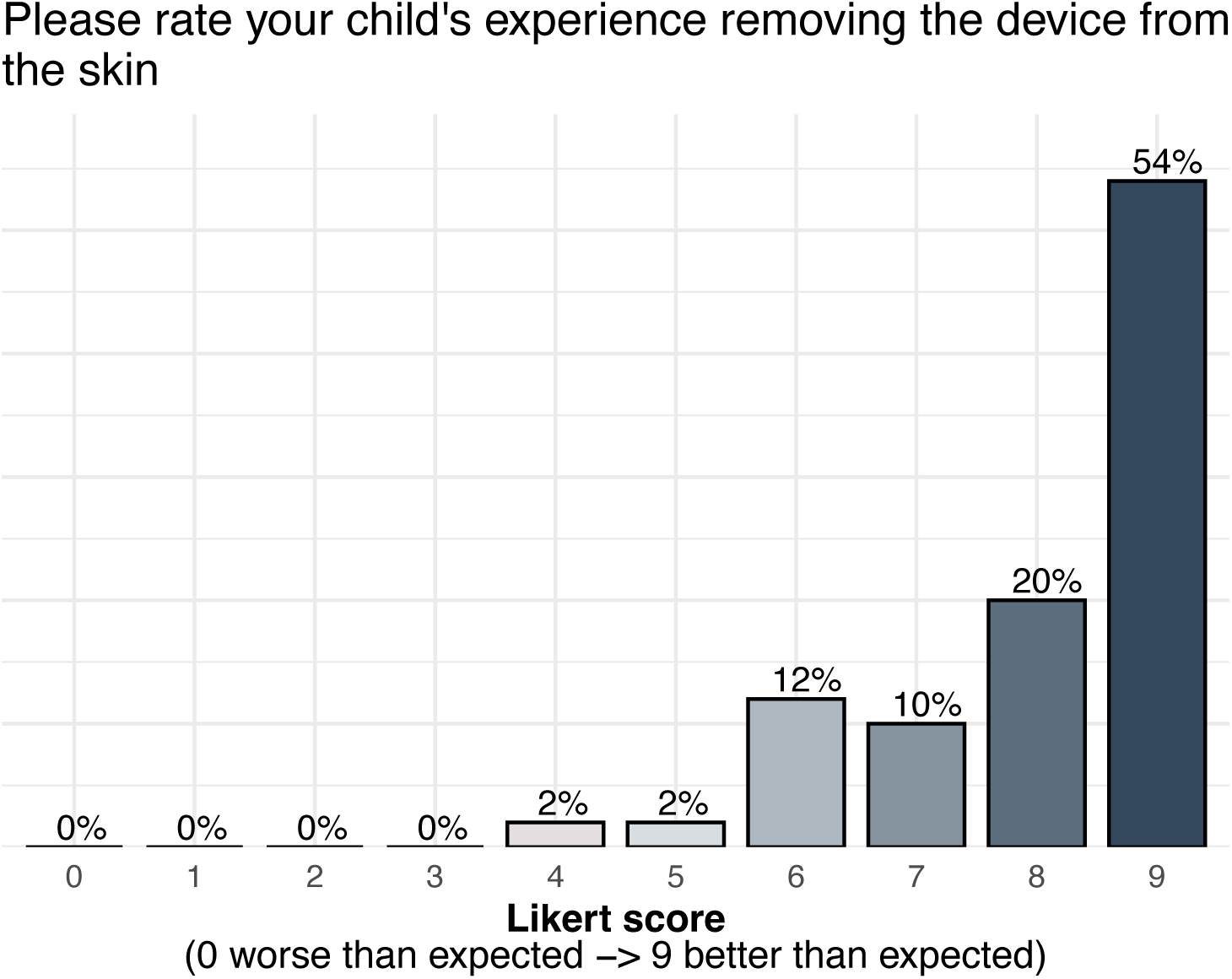
Parent-reported child experience during device removal. Bar chart showing parent-reported ratings of their child’s experience during removal of the RedDrop ONE device from the skin. Experience was assessed using scale, where 0 indicates worse than expected and 9 indicates better than expected. Bars represent the percentage of responses at each score. Ratings were predominantly clustered at the higher end of the scale, indicating that device removal was generally well tolerated.

**Figure 15.**
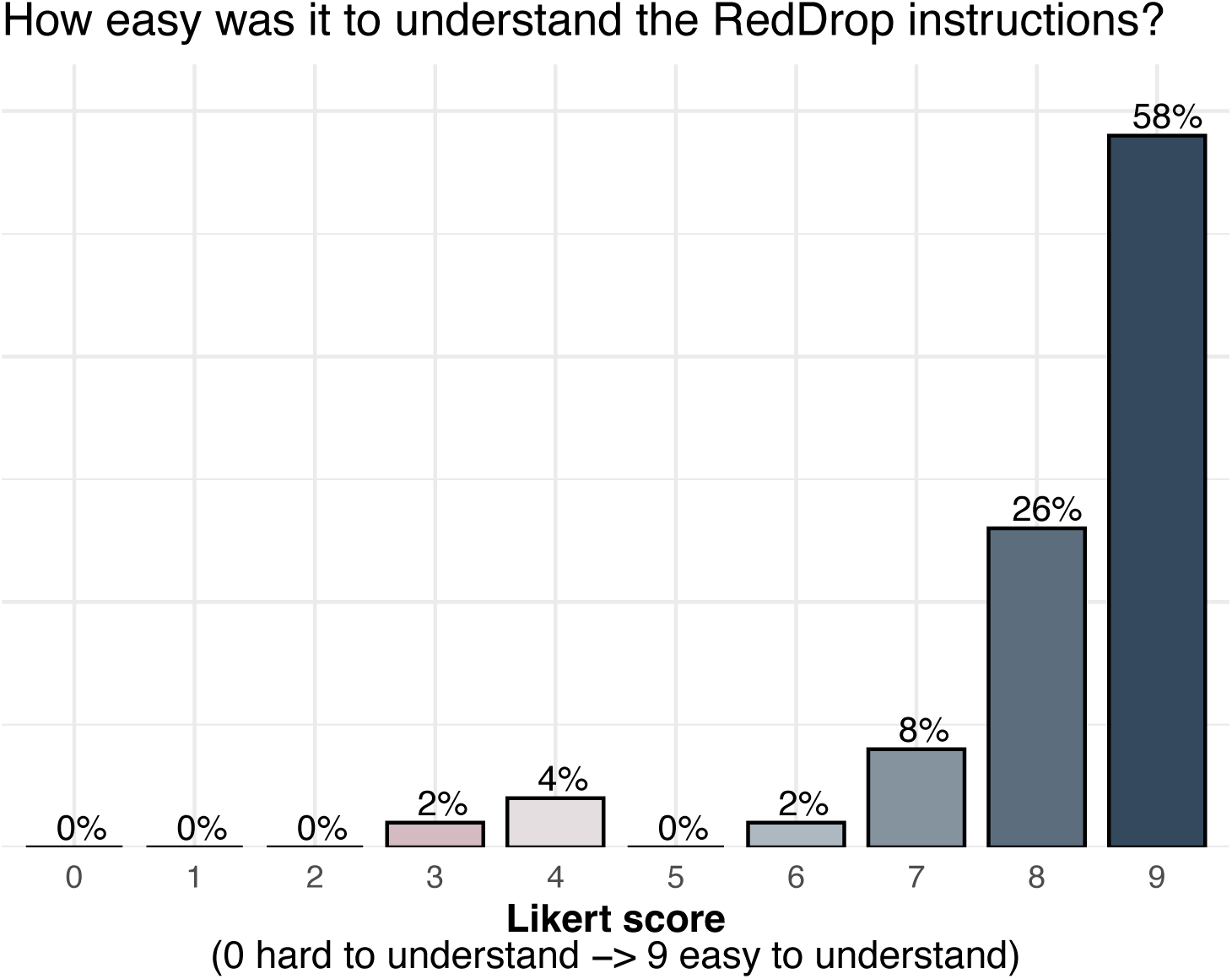
Parent-reported ease of understanding RedDrop ONE instructions. Bar chart showing parent-reported ratings of how easy the RedDrop ONE instructions were to understand. Ease of understanding was assessed using a scale, where 0 indicates hard to understand and 9 indicates easy to understand. Bars represent the percentage of responses at each score. Responses were predominantly distributed toward the higher end of the scale, indicating that most parents found the instructions easy to understand.

### Children responses

A subset of enrolled children elected to complete child-reported experience questions following blood collection. Of the total cohort, 36 children provided self-reported responses, while 14 children did not participate in the child questionnaire, reflecting the optional nature of child reporting. Among children who responded, reported pain levels were low for both collection sites. On a numeric rating scale from 0 (no pain) to 9 (most pain), arm-based pain scores had an average of 1.78 and a median of 1.00, while leg-based pain scores had an average of 1.61 and a median of 1.00 (**Figure 16 a-b**). These values indicate minimal perceived pain from the child’s perspective across both sites, with distributions clustered toward the lower end of the scale. Children also reported positive overall experiences with the blood collection process. Overall experience ratings, reflecting perceptions before, during, and after collection, had an average score of 7.94 and a median score of 8.00, indicating a generally favorable experience (**Figure 17**). Higher experience scores were common, while lower ratings were infrequent and did not show a consistent association with higher pain scores at either site. Together, these findings demonstrate that children who were able and willing to self-report experienced low levels of pain and generally positive overall experiences during parent-administered blood collection using the RedDrop ONE device. These child-reported outcomes further support the acceptability of the device and complement parent-reported usability and pain assessments in pediatric home settings.

**Figure 16.**
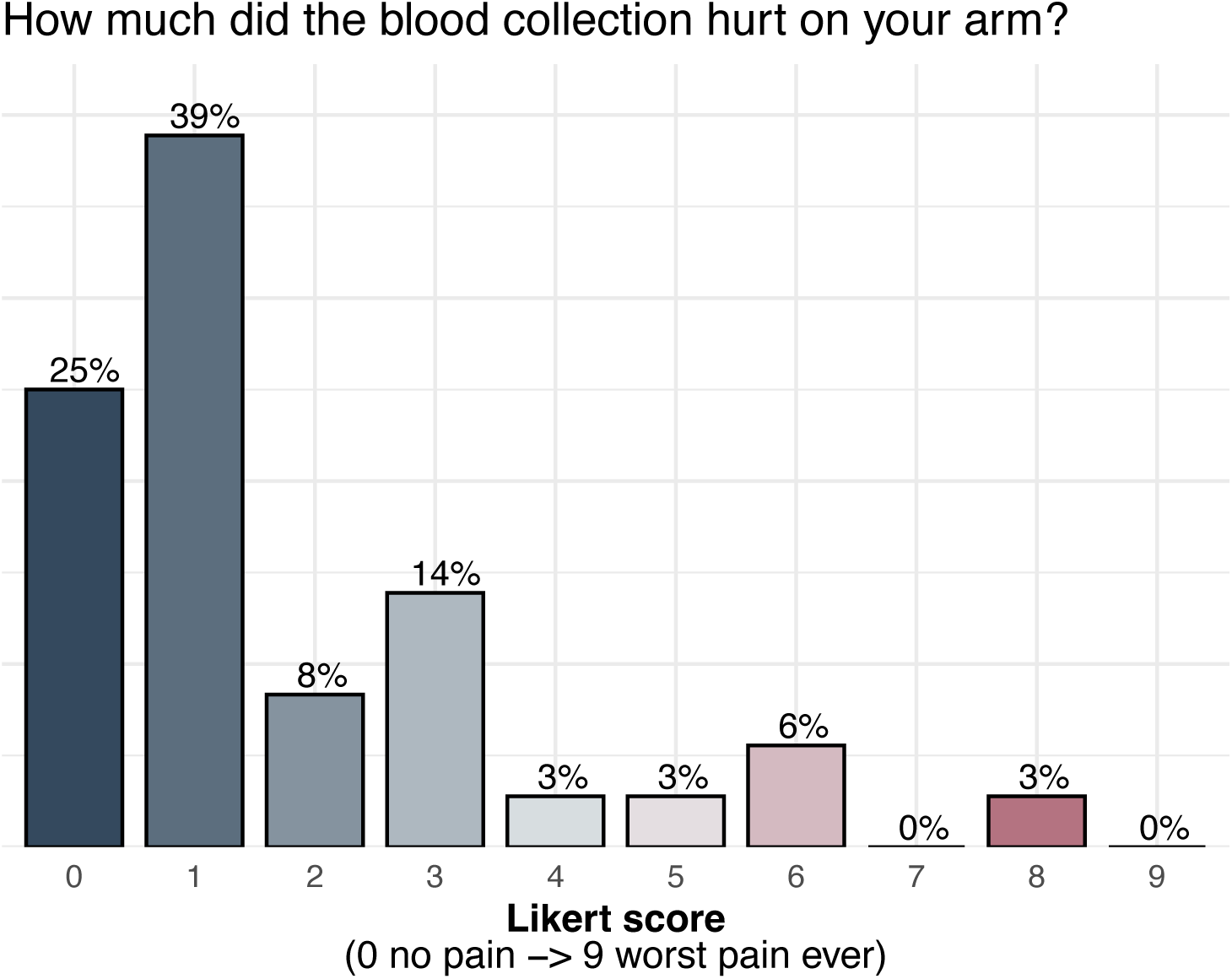
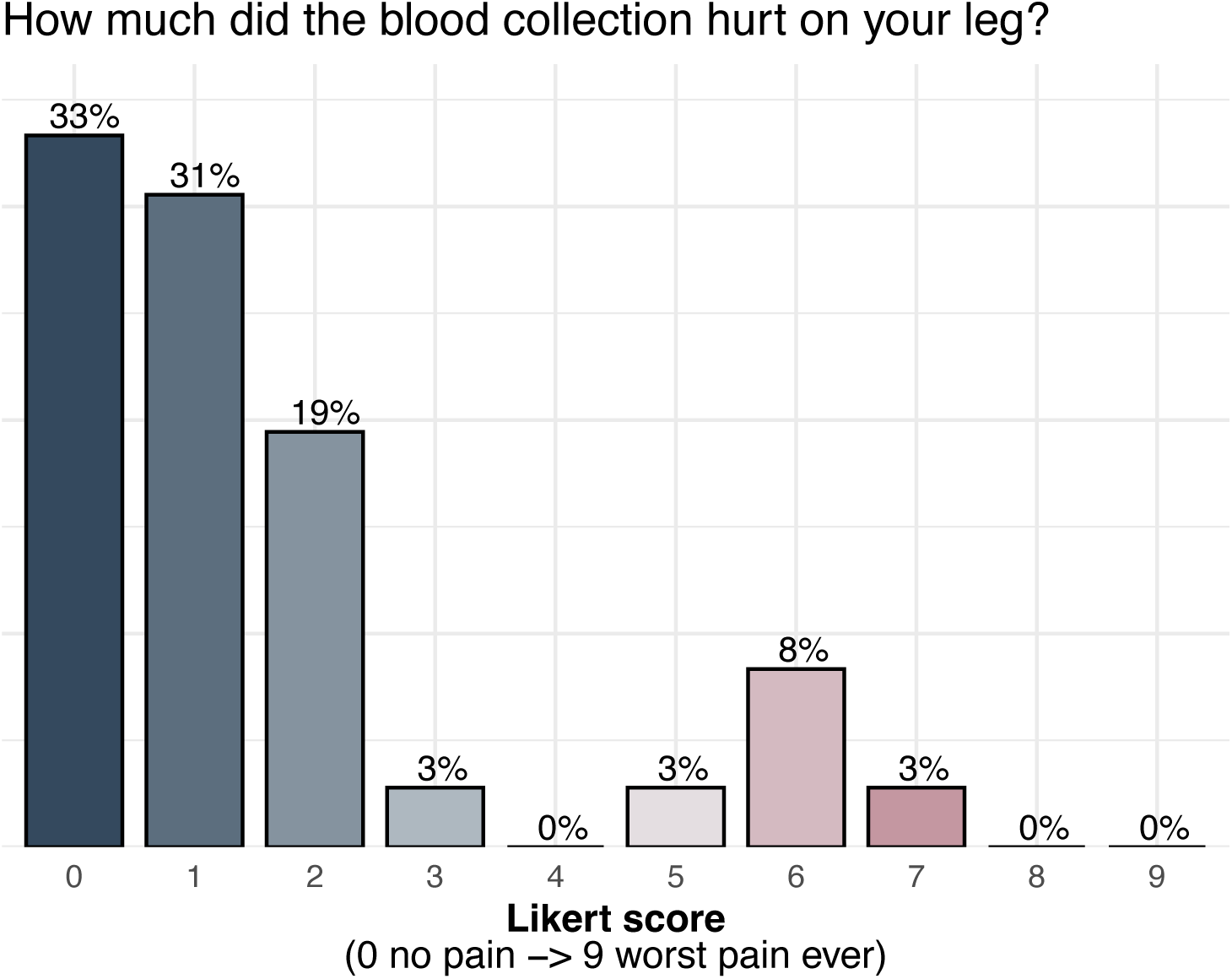
Child-reported pain associated with RedDrop ONE blood collection by anatomical site. **(a)** Distribution of child-reported pain scores for arm-based RedDrop ONE blood collection. **(b)** Distribution of child-reported pain scores for leg-based RedDrop ONE blood collection. Pain was assessed using a scale, where 0 indicates no pain and 9 indicates the worst pain ever experienced. Bars represent the percentage of responses at each pain score. Across both anatomical sites, pain ratings were predominantly clustered at the lower end of the scale, indicating low perceived pain associated with RedDrop ONE blood collection from the child’s perspective.

**Figure 17.**
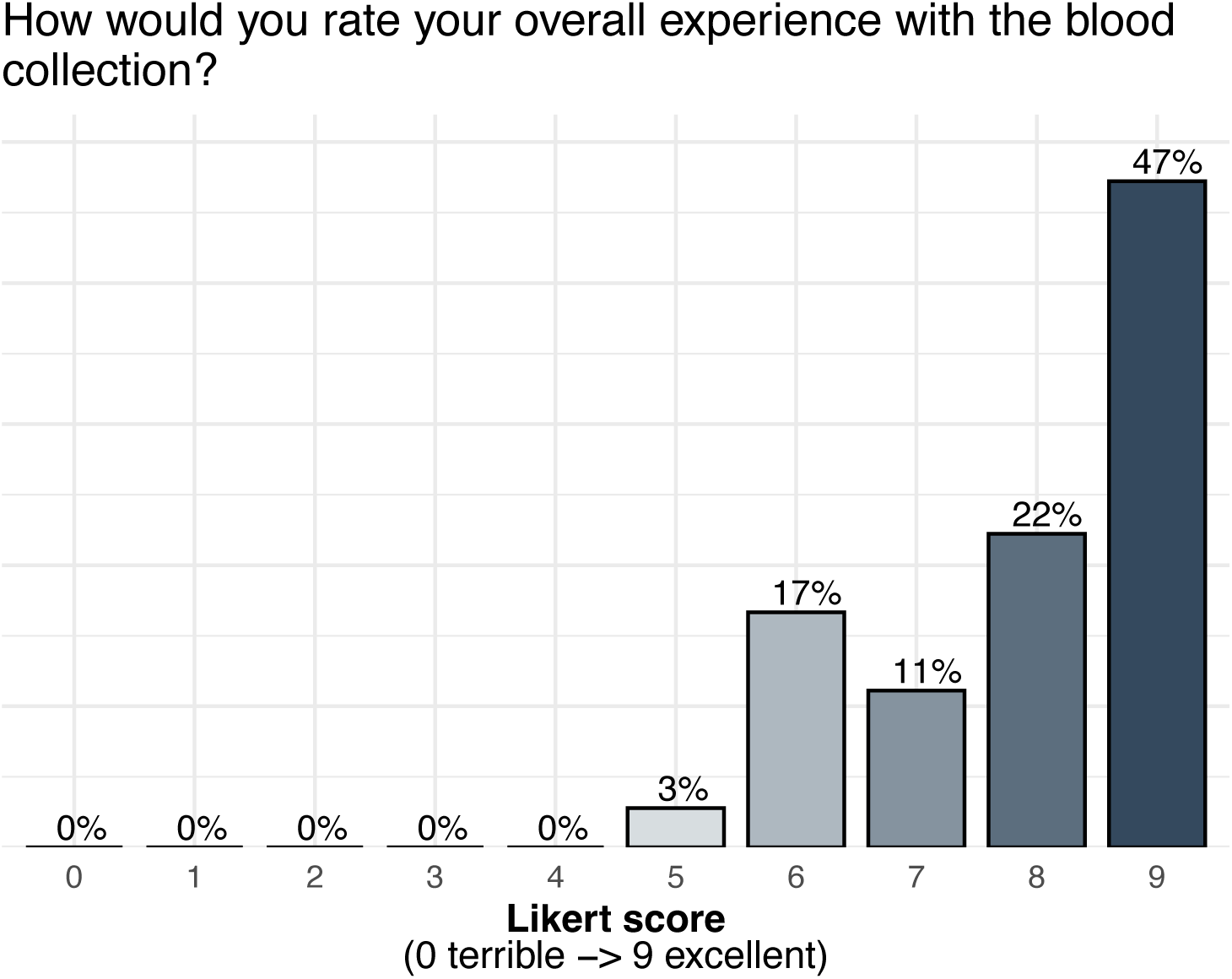
Child-reported overall experience with RedDrop ONE blood collection. Bar chart showing child-reported ratings of overall experience with RedDrop ONE blood collection. Experience was assessed using a scale, where 0 indicates a terrible experience and 9 indicates an excellent experience. Bars represent the percentage of responses at each score. Ratings were predominantly distributed toward the higher end of the scale, indicating a generally positive overall blood collection experience from the child’s perspective.

### Lipid Test

Paired lipid measurements obtained from arm and leg capillary samples demonstrated strong concordance across all analytes evaluated, including HDL, triglycerides, total cholesterol, and LDL. Distributions of values were highly overlapping between collection sites, with consistent central tendencies and comparable variability observed for each lipid parameter (**Figure 18**). These findings indicate a high degree of correlation between arm- and leg-derived samples collected during the same session, supporting analytical equivalence across alternative capillary collection sites. The observed consistency confirms that leg-based sampling yields lipid measurements comparable to those obtained from the arm, reinforcing flexibility in collection site selection for pediatric at-home blood sampling.

**Figure 18.**
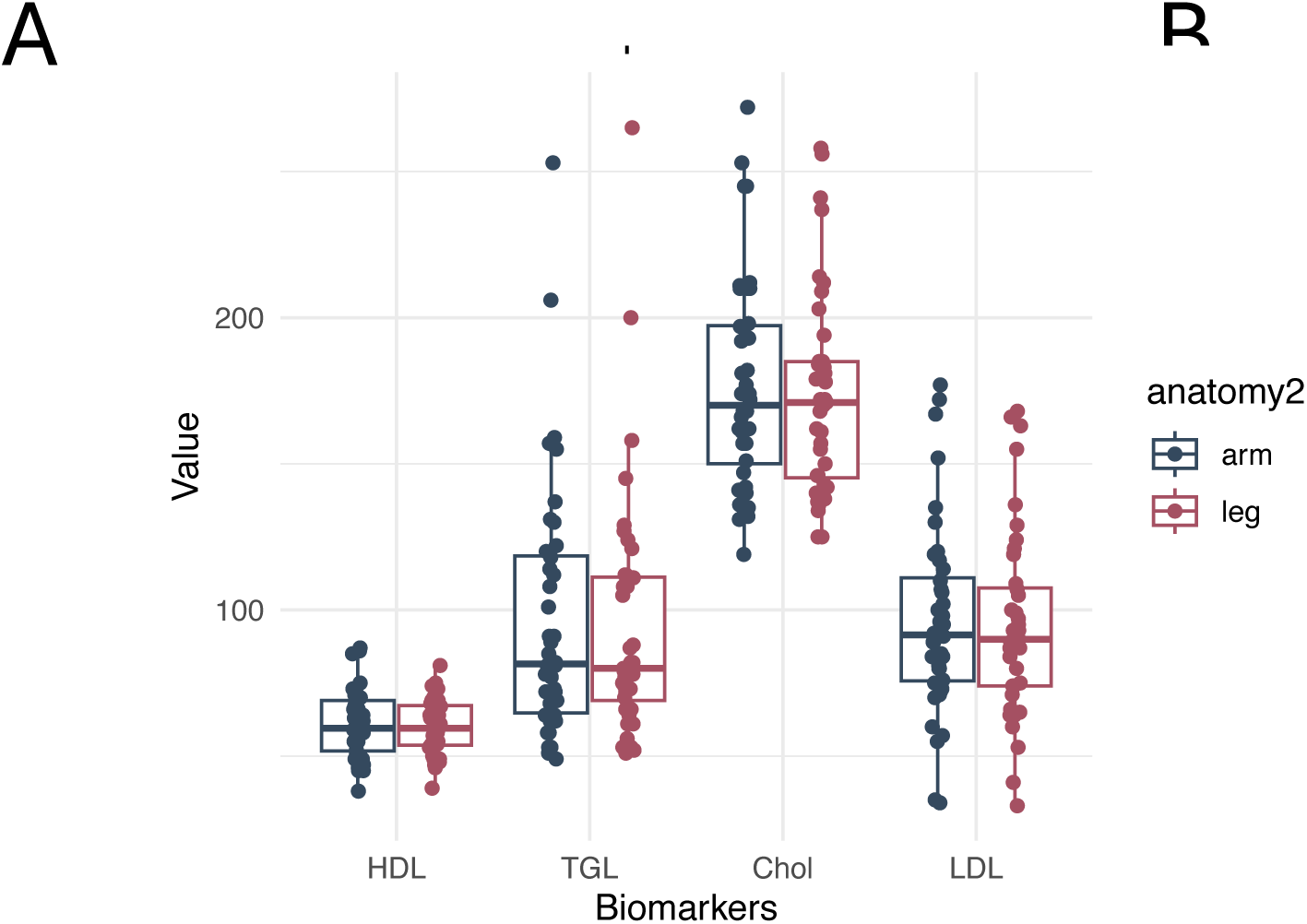
Lipid biomarker measurements from parent-administered capillary blood collection by anatomical site. Box-and-jitter plots showing lipid biomarker values measured from capillary blood samples collected using the RedDrop ONE device from arm and leg sites. Biomarkers include high-density lipoprotein (HDL), triglycerides (TGL), total cholesterol (Chol), and low-density lipoprotein (LDL). Each point represents an individual sample; box plots indicate the median and interquartile range. Comparable distributions were observed between arm and leg sampling sites across lipid biomarkers, indicating consistency of analyte measurements obtained from multiple anatomical collection locations under parent-administered, at-home conditions.

Paired lipid measurements obtained from arm and leg capillary samples demonstrated strong linear relationships across all analytes evaluated, including high-density lipoprotein (HDL), low-density lipoprotein (LDL), total cholesterol, and triglycerides (**Figure 19**). For each lipid parameter, values derived from leg samples increased proportionally with corresponding arm-derived measurements, with data points closely aligned along the regression lines across the full analytical range. The tight clustering of observations and the absence of systematic deviation at lower or higher concentration ranges indicate minimal site-dependent bias for all lipid analytes. Variability between paired arm and leg measurements was limited and consistent across HDL, LDL, total cholesterol, and triglycerides, supporting high concordance between collection sites.

**Figure 19.**
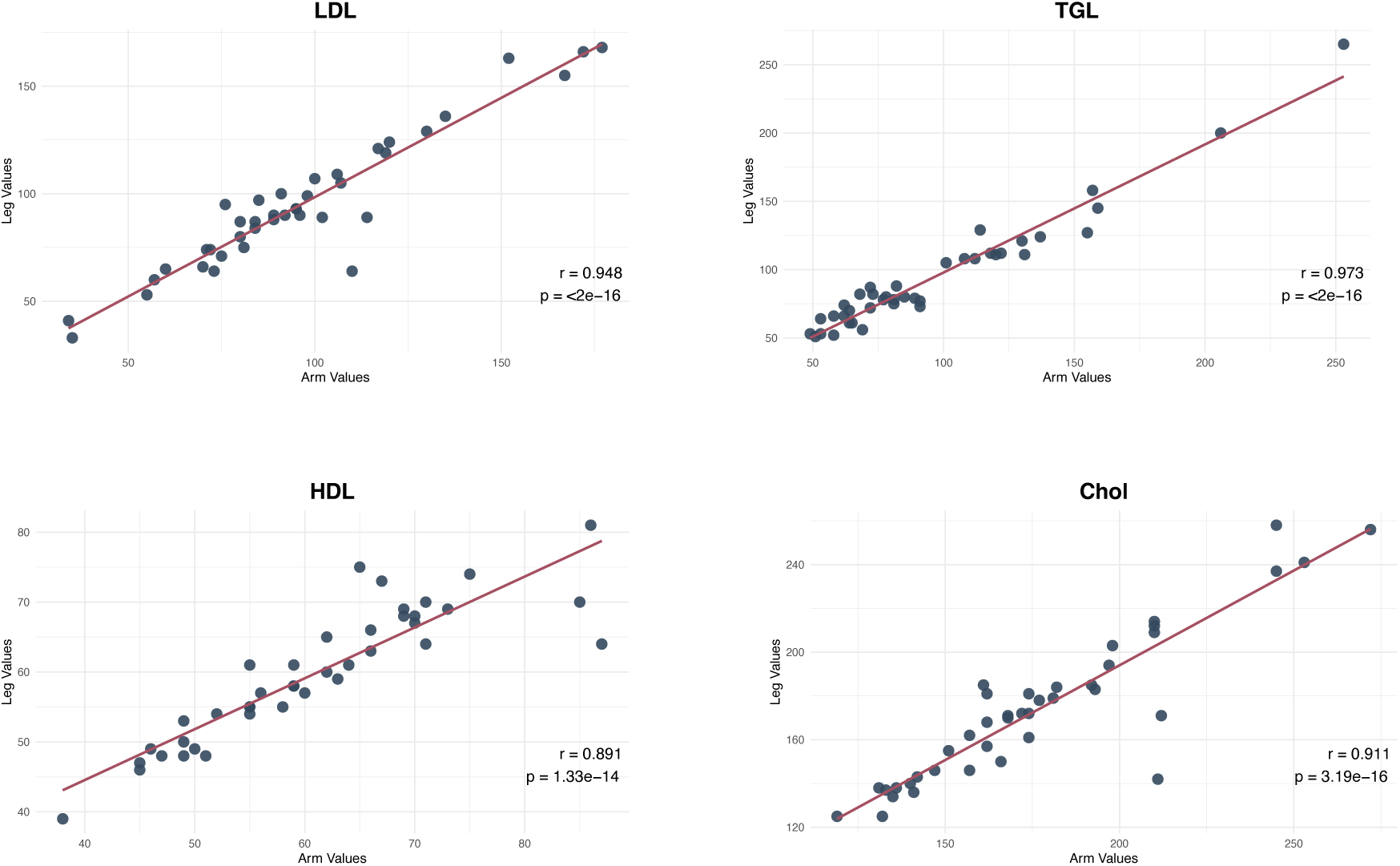
Correlation of lipid biomarker measurements between arm- and leg-derived capillary blood samples. Scatter plots showing pairwise correlations between lipid biomarker values measured from arm- and leg-collected capillary blood samples obtained using the RedDrop ONE device. Biomarkers include low-density lipoprotein (LDL), triglycerides (TGL), high-density lipoprotein (HDL), and total cholesterol (Chol). Each point represents an individual participant with paired arm and leg measurements. Solid lines indicate linear regression fits. Pearson correlation coefficients (*r*) and corresponding *p*-values are shown for each biomarker, demonstrating strong concordance between anatomical sampling sites under parent-administered, at-home collection conditions.

Collectively, these findings confirm that leg-based capillary sampling using the RedDrop ONE device yields lipid measurements comparable to those obtained from the arm when collected during the same session, supporting analytical equivalence across alternative capillary sampling sites and reinforcing flexibility in site selection for parent-administered pediatric lipid testing.

### Parent-Reported Fill Level as a Predictor of Blood and Serum Yield

Parent-reported fill level at the time of collection showed strong agreement with laboratory-measured total blood volume upon receipt (**Figure 20**). For both leg and arm samples, median measured blood volumes increased in a graded and predictable manner across parent-identified fill categories (first line, second line, above the second line, and whole tube), indicating close correspondence between parental observation and objective volume measurement. Across all fill levels, distributions of measured blood volume were well separated with limited overlap between adjacent categories, demonstrating that parents were able to accurately assess the extent of tube fill during at-home collection. This relationship was consistent for both leg and arm collections, with similar variability patterns observed within each fill category.

**Figure 20.**
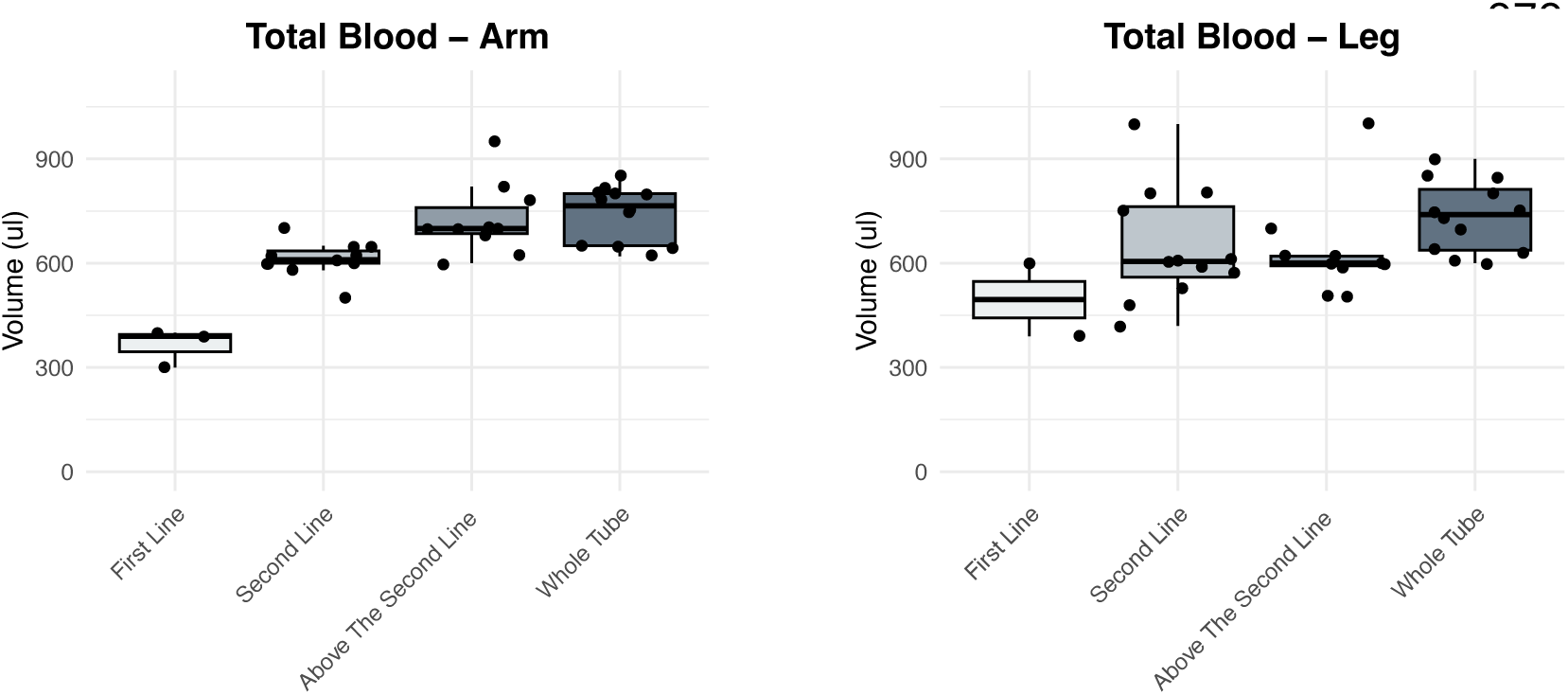
Parent-perceived tube fill thresholds in relation to measured whole blood volume by anatomical site. Box-and-jitter plots showing the relationship between parent-perceived tube fill thresholds and the corresponding measured total whole blood volumes collected using the RedDrop ONE device from the arm (left) and leg (right). Parents categorized collections based on visual fill thresholds (First Line, Second Line, Above the Second Line, and Whole Tube) at the time of sampling. Each point represents an individual collection, and box plots indicate the median and interquartile range of analytically measured blood volume at the Lab. Measured volumes increased systematically across perceived fill categories for both the arm and leg sites, demonstrating concordance between parent-assessed visual fill thresholds and quantitative blood volume.

These findings indicate that parent-reported fill level is a reliable proxy for actual collected blood volume and aligns closely with laboratory quantification. The strong concordance between parental observation and measured volume supports the feasibility and reliability of parent-administered capillary blood collection using the RedDrop ONE device, as well as the accuracy of at-home procedural guidance provided to families.

## Discussion

This RedDrop ONE study validates parent-administered pediatric capillary blood collection as a feasible, reliable, and scalable approach for use in real-world home settings. Across a geographically and clinically diverse cohort, parents successfully collected clinically meaningful blood and serum volumes from children using the RedDrop ONE device, with high acceptance, low reported pain, minimal device failure, and no adverse events. These findings support the premise that pediatric blood collection can be safely decentralized from traditional clinical environments without compromising sample quality or analytical performance.

Experience Survey data further strengthen this conclusion by demonstrating that usability and acceptability extend beyond successful sample acquisition. Parents reported high confidence in using the device, minimal difficulty during the collection process, and a high likelihood of completing collections on the first attempt. Most collections were completed efficiently, with a short overall time burden, reinforcing the practicality of integrating at-home blood collection into daily family routines without introducing meaningful disruption.

A central finding of this study is the demonstrated equivalence between arm and leg capillary collection sites. Blood and serum volumes collected from both sampling sites were comparable, with overlapping distributions, similar central tendencies, and narrow interquartile ranges, indicating consistent performance across collection sites. Lipid analyses, including HDL, LDL, total cholesterol, and triglycerides, showed strong concordance and linear correlation between paired arm and leg samples collected during the same session, with no evidence of site-dependent bias across the analytical range. Together, these results confirm that alternative pediatric capillary collection sites can be used interchangeably, providing flexibility in site selection based on child comfort, anatomy, or the need for multiple samples within a single session.

Experience Survey results also reinforce this flexibility. Parents reported no strong preference for arm or leg collection, with the largest proportion indicating no difference in ease of use between sites. Where preferences were expressed, they were relatively evenly distributed. Pain ratings were low for both sites, with leg-based collection perceived as slightly less painful. These findings suggest that site selection can be guided by child preference, comfort, or logistical considerations rather than technical constraints, an important feature for repeated or longitudinal sampling.

Importantly, this study also demonstrates that parents are capable of accurately assessing collection success in the home. Parent-reported sample tube fill level closely aligned with laboratory-measured blood volumes, with clear separation between fill categories and predictable increases in measured volume across reported thresholds. This concordance indicates that parental observation is a reliable proxy for actual sample yield and supports the effectiveness of the instructional materials, digital guidance, and survey workflows provided through the O’Ryan Health Parent Portal.

Reliable parent-reported metrics are a critical enabling feature for scalable home-based pediatric diagnostics and longitudinal monitoring programs.

Experience data further support parental competence and comfort with the process. Parents reported high confidence scores overall, even in cases where minor issues were encountered, suggesting that small challenges did not meaningfully interfere with successful collection. In addition, visible child discomfort was reported infrequently and was not associated with failure to complete collection or increased collection time, indicating that the procedure was well tolerated in real-world home settings.

Beyond device performance, the results underscore the importance of system-level infrastructure in enabling pediatric at-home blood collection. By embedding RedDrop ONE within the O’Ryan Health Artemis Platform, this study evaluated not only the usability of the device but also the feasibility of national deployment, virtual enrollment, compliant consent, guided collection, and overnight ambient-temperature shipping.

Families engaged with the study across 25 U.S. states, sampling in 19 states, including geographically remote regions, demonstrating that pediatric home blood collection can be operationalized at scale through an integrated logistics and informatics platform. This end-to-end capability helps differentiate RedDrop ONE as part of a repeatable at-home pediatric solution rather than a standalone collection tool.

The experience results highlight how this infrastructure directly contributes to the quality of life. Parents reported favorable comparisons between RedDrop ONE and traditional blood collection methods, rating the at-home experience as better than both finger prick and needle-based blood draws. High overall experience scores from both parents and children indicate that the combination of device design, instructional support, and platform-guided workflows reduces anxiety and burden associated with at-home pediatric blood collection.

The blood and serum volumes achieved in this study are sufficient to support routine clinical testing and, importantly, exceed thresholds commonly required for more advanced downstream molecular applications. While this study focused on lipid analysis as a representative clinical use case, the observed volumes and consistency support future use in metabolic, proteomic, transcriptomic, and genomic workflows. The ability to collect adequate volume from children at-home expands the potential for remote monitoring, longitudinal sampling, and participation in research and care pathways that would otherwise require repeated clinic visits. From a family perspective, minimizing repeated clinic and laboratory visits represents a meaningful improvement in daily life, particularly for children with chronic or complex medical conditions. The ability to perform blood collection at-home reduces travel, scheduling challenges, and procedural stress while enabling consistent monitoring and research participation.

Collier et al., in a comparative study conducted by Labcorp, the “Labcorp Study”, evaluated multiple capillary blood collection devices, including RedDrop ONE, Tasso+®, TAP MicroSelect®, Comfort Draw®, and traditional fingerstick methods, under professionally collected conditions at Labcorp facilities. (Collier et al., 2025) This laboratory-based evaluation provided an important benchmark for device performance under controlled clinical conditions. The RedDrop ONE study herein extends this body of work by assessing RedDrop ONE under parent-administered, at-home conditions, addressing a key gap identified in the “Labcorp Study”. Within the “Labcorp Study”, device performance was assessed across multiple dimensions, including sample volume and analytical suitability. In a head-to-head comparison of capillary blood collection technologies conducted by Collier et al., the RedDrop ONE device produced the highest mean whole blood (approximately 601 µL) and serum volumes (approximately 289 µL) among devices evaluated, a finding attributed to the higher vacuum pressure generated by the device, while remaining within acceptable analytical performance parameters for most clinical assays. In comparison, our virtual, at-home, parent-led RedDrop ONE study demonstrated higher mean volumes: 639 µL whole blood and 341 µL serum. These findings suggest that the higher-volume performance observed under professional collection conditions is preserved when RedDrop ONE is deployed in a fully remote, parent-led setting. Beyond volume, analytical reliability was also evaluated. In the “Labcorp Study” serum obtained using the RedDrop ONE device demonstrated strong concordance with matched venous serum across a broad panel of routine clinical chemistry analytes. Consistent with these findings, lipid measurements obtained in our RedDrop ONE study showed tightly clustered distributions across HDL, triglycerides, total cholesterol, and LDL, with similar values observed between arm and leg collections, supporting the analytical consistency of RedDrop ONE-derived serum across routine lipid analytes. Together, these results indicate that RedDrop ONE supports both adequate sample volume and analytically consistent serum measurements across professional and fully remote, family-centered use contexts.

Schuchardt et al. conducted a cross-sectional “General Practice” study in two rural general practice clinics in Germany to evaluate the pain, feasibility, usability, and blood volume yield of Tasso+® capillary self-blood collection for upper-arm sampling in adult primary care patients (Schuchardt et al., 2025). In the “General Practice” study, participants performed first-time capillary self-blood collection independently in a clinical setting, with 57.5% (61/106) completing the procedure without assistance. In our virtual, at-home, parent-led study, 100% of parents completed capillary blood collection without any medical assistance, demonstrating complete sampling success under fully remote conditions, with instruction provided solely through video and photo-based materials. In the “General Practice” study, a predefined threshold of 130 µL plasma was used for routine laboratory testing, 59.4% of participants (63/106) met or exceeded this volume. In contrast, in the RedDrop ONE study, all collections were completed without medical assistance, and all participants successfully obtained substantially higher blood and serum volumes. In the RedDrop ONE study, the median total blood volume was 615 µL, and the median serum volume was 340 µL. Notably, the lowest observed serum volumes were 150 µL for arm collections and 180 µL for leg collections, exceeding the predefined threshold of 130 µL volume in the “General Practice” study. The RedDrop ONE Study findings indicate consistently higher collection success and sample adequacy under fully parent-administered conditions compared the “General Practice” study designed as a self-collection experience in clinical settings.

Recent work has demonstrated the feasibility of fully decentralized blood collection at a national scale in the U.S. In the “homeRNA study”, Lim et al. reported that 68 adult participants across 26 U.S. states completed a fully remote, at-home self-collection protocol using Tasso+®-SST and an RNA stabilization tube, the homeRNA platform, returning 691 longitudinal blood samples by standard mail under ambient shipping conditions with variable real-world temperature exposure over approximately one year (Lim et al., 2025). The “homeRNA study” demonstrated that direct-to-home kit distribution, participant-led self-collection, and mail-based return logistics can support large-scale remote biospecimen collection across diverse geographic regions. Although developed independently, the design and outcomes of the present RedDrop ONE study are consistent with these findings and provide complementary evidence in a distinct context. Specifically, The RedDrop ONE study applies a fully remote, parent-led model in a pediatric population using RedDrop ONE device and focuses on blood and serum analytics rather than RNA stabilization. A key distinction is that the “homeRNA study” enrolled adults aged 18 years and older (median age approximately 36 years), whereas the RedDrop ONE study evaluates parent-led blood collection in children ranging from 3-17 (median age 13 years), extending decentralized sampling into a younger and more vulnerable population. Both studies share key operational features, including decentralized enrollment, electronic consent, direct-to-home kit distribution, remote instruction, and mail-based sample return without clinical supervision. Notably, our results demonstrate that parents can successfully perform capillary blood collection at-home using only video- and photo-based instructions to reliably obtain sufficient blood and serum volumes for routine clinical laboratory testing, supporting the feasibility of extending decentralized blood collection into family-centered pediatric care.

Several limitations warrant consideration. This was a usability and feasibility study designed for descriptive evaluation rather than hypothesis testing, and blood analysis was conducted at a single CLIA-certified laboratory. Pre-existing clinical conditions were parent-reported, and the study did not include venous comparator samples. These limitations are appropriate for the study’s objectives. In the study’s favor, all primary data generation in this study, including blood collection and laboratory analysis, was performed independently of O’Ryan Health: samples were collected by parents in the home and analyzed by an external CLIA-certified laboratory, with O’Ryan Health serving solely as the coordinating entity responsible for study logistics, data aggregation, and descriptive reporting. This separation of roles reduces sponsor- or operator-driven bias and reflects real-world deployment conditions for decentralized pediatric testing.

In conclusion, the RedDrop ONE study demonstrates that parent-administered pediatric blood collection using the RedDrop ONE device can be successfully implemented in the home, expanding access to decentralized blood testing for children. Data demonstrates equivalent performance across multiple home settings, accurate parent-reported collection metrics, reliable collected blood volumes for analysis, and successful national deployment through the O’Ryan Health Artemis Platform. The RedDrop ONE study supports a new model for pediatric blood collection that minimizes home life disruption from repeated clinic and laboratory visits, meaningfully improves quality of life for children and parents, expands access to testing, and empowers parents to safely participate in their child’s care at home. The RedDrop ONE capillary blood collection device has the potential to transform pediatric research, remote monitoring, and routine care by bringing clinically meaningful blood collection into the home.

## Data Availability

All data produced in the present study are available upon reasonable request to the authors

## Acknowledgements

We extend our sincere gratitude to the parents and children who participated in this study for their trust, commitment, and courage in helping to pioneer new approaches to pediatric healthcare. We thank BRANY IRB for providing ethical review and regulatory oversight, and Zhiyi Qiang from SanoCardio for performing all clinical blood analyses. We acknowledge Debojyoti Singha and Ananya Singha of Swing Technologies for the development of Version 1 of the O’Ryan Health Artemis “Superhero Software Superstack”, which enabled virtual study operations. We also thank Vernice Angelica Lat and Junessa Masaya for developing educational and marketing content supporting study engagement, and Biomedical Reasoning, LLC, for assistance with data processing and analytical methodology.

---

| Survey Question | Scale | Average Rating | Median Rating | Standard Deviation |
| --- | --- | --- | --- | --- |
| Please rate your child's pain experienced from the RedDrop ONE on the ARM. | 0 = no pain,<br>9 = worst pain ever | 2.32 | 2.00 | 2.10 |
| Please rate your child's pain experienced from the RedDrop ONE on the LEG. | 0 = no pain,<br>9 = worst pain ever | 2.34 | 1.50 | 2.33 |
| Please rate your child's experience using the heat pack. | 0 = no pain,<br>9 = worst pain ever | 1.28 | 0.00 | 2.38 |
| Please rate your experience activating the device, specifically the slider. | 0 = very difficult,<br>9 = very easy | 8.28 | 9.00 | 2.05 |
| Please rate your child's experience removing the device from the skin. | 0 = worse than expected,<br>9 = better than expected | 8.06 | 9.00 | 1.27 |
| Please rate your experience attaching the Microtainer tube from the device. | 0 = worse than expected,<br>9 = better than expected | 7.86 | 9.00 | 1.60 |
| Please rate your experience removing the Microtainer tube from the device | 0 = worse than expected,<br>9 = better than expected | 8.10 | 9.00 | 1.30 |
| Please rate your child's overall experience with the RedDrop ONE. | 0 = worse than expected,<br>9 = better than expected | 8.26 | 9.00 | 1.00 |
| How confident did you feel using the device on your child? | 0 = not confident,<br>9 = extremely confident | 8.50 | 9.00 | 0.94 |
| How long did the blood collection process take from start to finish? | minutes | 10.03 | 8.50 | 7.77 |
| How would you compare this device to a traditional finger prick? | 0 = much worse,<br>9 = much better | 7.92 | 9.00 | 1.52 |
| How would you compare this device to a traditional needle blood draw? | 0 = much worse,<br>9 = much better | 8.60 | 9.00 | 0.72 |
| How easy was it to understand the RedDrop ONE instructions? | 0 = hard to understand,<br>9 = easy to understand | 8.20 | 9.00 | 1.36 |
| Number of respondents, n=50 |  |  |  |  |

| Survey Question | Response: YES | Response: NO |
| --- | --- | --- |
| Were you able to successfully collect blood from the ARM site? | 50<br>(100%) | 0<br>(0%) |
| Did the device activate (click) properly on the ARM site? | 50<br>(100%) | 0<br>(0%) |
| Were you able to successfully collect blood from the LEG site? | 50<br>(100%) | 0<br>(0%) |
| Did the device activate (click) properly on the LEG site? | 50<br>(100%) | 0<br>(0%) |
| Did you notice any differences in your child's reaction between the ARM and LEG collection? | 24<br>(48%) | 26<br>(52%) |
| Were there any issues or difficulties during the collection process? | 10<br>(20%) | 40<br>(80%) |
| Were you able to complete the blood collection on your child on the first attempt? | 47<br>(94%) | 3<br>(6%) |
| Was there any visible discomfort in your child during or after collection? | 13<br>(26%) | 37<br>(74%) |
| Would you be willing to use RedDrop ONE device again for at-home blood collection on your child? | 49<br>(98%) | 1<br>(2%) |
| Did you need any assistance beyond the provided instructions? | 3<br>(6%) | 47<br>(94%) |
| Would you be willing to participate in a future study using the RedDrop ONE? | YES: 41<br>(82%)<br>MAYBE: 7<br>(14%) | 2<br>(4%) |
| Does your child want to answer some questions? | 36<br>(72%) | 14<br>(28%) |
| Number of respondents, n=50 |  |  |

| Survey Question | Scale | Average Rating | Median Rating | Standard Deviation |
| --- | --- | --- | --- | --- |
| **For children:**<br>**How much did the blood collection hurt on your ARM?** | 0 = no pain,<br>9 = worst pain ever | 1.78 | 1.00 | 1.96 |
| **For children:**<br>**How much did the blood collection hurt on your LEG?** | 0 = no pain,<br>9 = worst pain ever | 1.61 | 1.00 | 1.98 |
| **For children:**<br>**How would you rate your overall experience with the blood collection (before, during, and after)?** | 0 = terrible,<br>9 = excellent | 7.94 | 8.00 | 1.24 |
| Number of respondents, n=36 |  |  |  |  |

| Survey Question | Response: YES | Response: NO |
| --- | --- | --- |
| For Children:<br>Were you scared before using the device? | 21<br>(58%) | 15<br>(42%) |
| For Children:<br>Would you be okay using the device again in the future? | 36<br>(100%) | 0<br>(0%) |
| For Children:<br>Would you be okay with using the RedDrop ONE on your ARM again in the future? | 33<br>(92%) | 3<br>(8%) |
| For Children:<br>Would you be okay with using the RedDrop ONE on your LEG again in the future? | 35<br>(97%) | 1<br>(3%) |
| Number of respondents, n=36 |  |  |

| Survey Question | Response 1 | Response 2 | Response 3 | Response 4 |
| --- | --- | --- | --- | --- |
| <sup>1</sup> How much blood was collected from the ARM? | Whole tube: 18, (36%) | Above second line: 11, (22%) | Second line: 14, (28%) | First line: 7 (14%) |
| <sup>1</sup> How much blood was collected from the LEG? | Whole tube: 17, (34%) | Above second line: 11, (22%) | Second line: 16, (32%) | First line: 6 (12%) |
| <sup>1</sup> Which site was easier to use the RedDrop ONE device on? | Arm: 13, (26%) | No difference: 22, (44%) | Leg: 15, (30%) | n/a |
| <sup>2</sup> For Children: How did it feel during and after? | Didn't hurt at all: 10, (28%)<br>Didn't hurt + initially nervous: 3, (8%) | Hurt a little but was ok: 14, (39%)<br>Hurt a little + initially nervous: 4, (11%) | Hurt a lot: 0, (0%) | Was nervous but it was fine: 5, (14%)<br>Total feeling nervous: 12, (33%) |
| <sup>1</sup> Number of respondents, n=50, <sup>2</sup> Number of respondents, n=36 |  |  |  |  |

## Footnotes

1 RedDrop ONE. *510(k) Summary for RedDrop ONE (One).* U.S. Food and Drug Administration. 510(k) Number: K234081. Available at: https://www.accessdata.fda.gov/cdrh_docs/pdf23/K234081.pdf.

2 RedDrop ONE. RedDrop One – Personal Blood Lancet Device. 2024. Product overview and specifications sheet.

3 RedDrop ONE. RedDrop ONE Clinical Trial Report. Document No. TR-25-0021, Revision X1. December 15, 2023.

4 The OFF-score is part of the Athlete Biological Passport (ABP), particularly the hematological module, which monitors blood variables over time rather than relying on a single test.

## References

Collier, B.B., Brandon, W.C., Chappell, M.R., Iacovetti, G., Tokunaga, N., Schaff, U.Y., Sommer, G.J., and Grant, R.P. (2025). Comparing capillary blood collection technologies: assessing patient experience, device performance, & clinical accuracy. Bioanalysis 17, 1329–1336.

Jebb, A.T., Ng, V., and Tay, L. (2021). A Review of Key Likert Scale Development Advances: 1995-2019. Front Psychol 12, 637547.

Lewis, L., Smith, M., Boutard, K., Fedoruk, M., and Miller, G. (2025). Comparison of Microcapillary Blood Sampling Devices for Use in Anti-Doping. Drug Test Anal 17, 1145–1149.

Lim, F.Y., Lea, H.G., Dostie, A.M., Kim, S.Y., van Neel, T.L., Hassan, G.W., Takezawa, M.G., Starita, L.M., Adams, K.N., Boeckh, M., et al. (2025). homeRNA self-blood collection enables high-frequency temporal profiling of presymptomatic host immune kinetics to respiratory viral infection: a prospective cohort study. EBioMedicine 112, 105531.

Mahendru, D., Kumaravel, J., Mahalmani, V.M., and Medhi, B. (2020). Athlete Biological Passport: Need and Challenges. Indian J Orthop 54, 264–270.

Schuchardt, C., Muller, F., Hafke, A., Hummers, E., Schanz, J., Dopfer-Jablonka, A., Behrens, G.M.N., and Schroder, D. (2025). Pain and feasibility of capillary self-blood collection in general practice: A cross-sectional investigative study. Eur J Gen Pract 31, 2501309.

